# Develop and Validate A Fair Machine Learning Model to Indentify Patients with High Care-Continuity in Electronic Health Records Data

**DOI:** 10.1101/2025.11.11.25339938

**Authors:** Yao An Lee, Tiange Tang, Yu Huang, Jiang Bian, Lizheng Shi, Jingchuan Guo

**Author notes:** The two authors contributed equally and serve as co-first authors (Yao An Lee, Tiange Tang). The two authors served as co-corresponding authors**, Corresponding authors**: Jingchuan Guo, MD, PhD, Affiliation: Department of Pharmacy Practice, Purdue University College of Pharmacy, Address: Fifth Third Bank Building at Eskenazi Health, 640 Eskenazi Ave, Indianapolis, IN 46202, Lizheng Shi, PhD, MsPharm, MA, Affiliation: Neal A. and Mary Vanselow Endowed Chair Director of Health Systems Analytics Research Center Professor of Department of Health Policy and Management, Celia Scott Weatherhead School of Public Health and Tropical Medicine, Tulane University Address: 1440 Canal Street, Suite 1900, New Orleans, LA 70112.

## Abstract

**Objectives:** Electronic health record (EHR) data often missed care outside a given health system, resulting in data discontinuity. We aimed to: (1) quantify misclassification across levels of EHR data discontinuity and identify an optimal continuity threshold. (2) develop a machine learning (ML) model to predict EHR continuity and optimize fairness across racial and ethnic groups, and (3) externally validate the EHR continuity prediction model using an independent dataset.

**Materials and Methods:** We used linked OneFlorida+ EHR–Medicaid claims data for model development and REACHnet EHR–Louisiana Blue Cross Blue Shield (LABlue) claims data for external validation. A novel Harmonized Encounter Proportion Score (HEPS) was applied to quantify patient-level EHR data continuity and the impact on misclassification of 42 clinical variables. ML models were trained using routinely available demographic, clinical, and healthcare utilization features derived from structured EHR data.

**Results:** Higher EHR data continuity was associated with lower rates of misclassification. A HEPS threshold of approximately 30% effectively distinguished patients with sufficient data continuity. ML models demonstrated strong performance in predicting high continuity (AUROC=0.77). Fairness assessments showed bias against Hispanic group, which was substantially improved following bias mitigation procedures. Model performance remained robust and fair in the external validation.

**Discussion:** Our study offers a practical metric for quantifying care continuity in EHR networks. The current ML model incorporating EHR-routinely collected information can accurately identify patients with high care continuity.

**Conclusions:** We developed a generalizable care-continuity classification tool that can be easily applied across EHR systems, strengthening the rigor of EHR-based research.

## INTRODUCTION

Electronic health records (EHRs) have become a cornerstone of real-world data (RWD), offering a wealth of clinical information that is increasingly utilized for research purposes. Among these studies, EHRs are particularly valuable in comparative effectiveness research (CER) because of their rich clinical detail, longitudinal structure, and growing accessibility through national research networks^1,2^. National network such as PCORnet, is a “network of networks” supported by the Patient-Centered Outcomes Research Institute (PCORI)^3,4^ and have enabled access to population-scale EHR data covering over 70 million patients across the U.S.^5^ However, a major limitation of EHR-based RWD is the fragmentation of the U.S. healthcare system, where patients often receive care across multiple, unlinked providers, and most EHR systems do not capture care delivered outside their affiliated settings. This discontinuity is further compounded by variability in healthcare access, driven in part by social determinants of health,^6,7^ which influence patients’ patterns of care-seeking across different systems. As a result, medical information recorded in other healthcare systems often remains “invisible” to investigators, leading to low EHR continuity. These challenges contribute to low EHR continuity, defined as “medical information recorded outside the reach of an EHR system,” posing a substantial risk of introducing information bias in research studies.^8,9^ Studies have shown that patients with low EHR continuity experience up to a 17-fold higher misclassification in key variables compared to those with high continuity^10^.

Without linkage to additional data sources, researchers face significant limitations in assessing the completeness of patient records, leaving their analyses vulnerable to potential misclassification of exposures, outcomes, and key covariates used in confounding control. In contrast, insurance claims data offer comprehensive capture of healthcare utilization within defined enrollment periods, although they often lack the clinical nuance present in EHRs.^11^ While combining EHRs with claims data can substantially enhance data completeness and reliability, such linkages remain uncommon due to challenges related to privacy regulations, data governance, and technical feasibility. These limitations highlight the pressing need to systematically measure EHR continuity and to establish scalable, transparent approaches for mitigating its influence on data integrity and the validity of real-world evidence generated from EHR-based studies. Patients with high EHR data continuity, where most clinical encounters are captured within a single EHR system, provide a more reliable foundation for clinical research and decision-making. Conversely, low EHR data continuity, characterized by fragmented records across disparate systems, can obscure important clinical information and exacerbate inequities in research and care.^6^

At the same time, while restricting CER cohorts to patients with high EHR continuity can improve validity, doing so without accounting for structural inequities may unintentionally exclude underserved populations, particularly racial and ethnic minorities with fewer care options.^12^ Evidence shows that machine learning (ML) systems trained on incomplete or biased data can perpetuate or amplify existing health disparities by underestimating care needs for Black patients when cost is used as a proxy for health status, or by failing to detect clinical signs in racial groups underrepresented in training datasets.^13^ As a result, there is increasing urgency to assess and ensure fairness in ML-driven healthcare tools. This includes evaluating algorithmic behavior using fairness metrics such as demographic parity, equalized odds, and predictive rate parity,^14,15,16,17^ all of which aim to prevent systematic disadvantages across protected subgroups. Despite the growing literature, there remains no consensus on which fairness metrics should be applied in different contexts, highlighting the need for careful selection and application based on the specific characteristics and implications of the healthcare setting.

## OBJECTIVE

The objective of this study was to improve the validity of EHR data and promote health equity by developing and validating a ML algorithm to identify patients with high EHR data continuity. Specifically, we (1) evaluated how EHR data discontinuity contributes to misclassification bias and identified an optimal HEPS threshold for acceptable data completeness in our EHR network; (2) developed a ML algorithm to predict EHR data continuity at the person level and assessed algorithmic fairness across racial and ethnic subgroups; and (3) externally validated the continuity prediction algorithm in an independent EHR dataset. Using linked EHR-Florida Medicaid claims data from OneFlorida+ network, we assessed the degree of misclassification across varying levels of data continuity to establish a threshold at which EHRs alone are considered sufficiently complete for valid inference. Furthermore, to ensure transparency and equity, we incorporated model explainability tool and applied fairness metrics to evaluate and mitigate disparities in model performance across racial and ethnic groups. Finally, we validated our continuity prediction algorithm in linked EHR–claims data from Research Action for Health Network (REACHnet) EHRs and Louisiana Blue (LABlue) claims network to evaluate generalizability and transportability. Collectively, this work sought to deliver a tool that improves research validity through rigorous identification of patients with adequate data continuity and guarantees equitable performance across heterogeneous populations and healthcare systems.

## MATERIALS AND METHODS

### Data Source

The study used data from (1) the OneFlorida+ EHR linked with Florida Medicaid claims data and (2) the REACHnet EHR linked with commercial claims data from Louisiana Blue (LABlue). Model development was conducted in linked OneFlorida+ EHR–Florida Medicaid claims data, and external validation was performed in the linked REACHnet–LABlue dataset. Both OneFlorida+ and REACHnet are members of the National Patient-Centered Clinical Research Network (PCORnet).^18^ Both datasets include rich, longitudinal EHRs and follow the PCORnet Common Data Model (CDM)^5^ to ensure consistency and interoperability. OneFlorida+ EHRs linked with Florida Medicaid claims data were used for algorithm development and internal validation, and REACHnet EHRs linked with commercial LABlue claims data were used for external validation. Claims data were used as gold standards to quantify the magnitude of data continuity on the EHR side.

### Study design and population

Using the OneFlorida+ clinical research network, we extracted patients with linked EHR and Florida Medicaid claims between January 2015 and October 2023. For external validation, we selected a parallel cohort with linked EHR–claims from the REACHnet–LABlue dataset between January 2022 and September 2024. Eligible patients from both networks met predefined criteria: (1) adults aged 18 or older, (2) had at least one encounter recorded in the EHR system during their active enrollment period. We excluded individuals who (1) had fewer than 12 months of EHR or claims data, or (2) had a secondary payer.

For each included patient, the date of the first recorded instance of linked EHR–claims data within the study period was designated as the index date (**Figure 1**), and patients were followed for one year thereafter to evaluate data continuity. In OneFlorida+, patients with a first complete year of data between 2015–2019 were used as the training/internal-validation cohort, whereas the cohort identified during 2020–2023 was used for independent testing. In the REACHnet–LABlue data, patients with a first complete year of data during 2022–2024 were used as the external-validation cohort.

**Figure 1.**
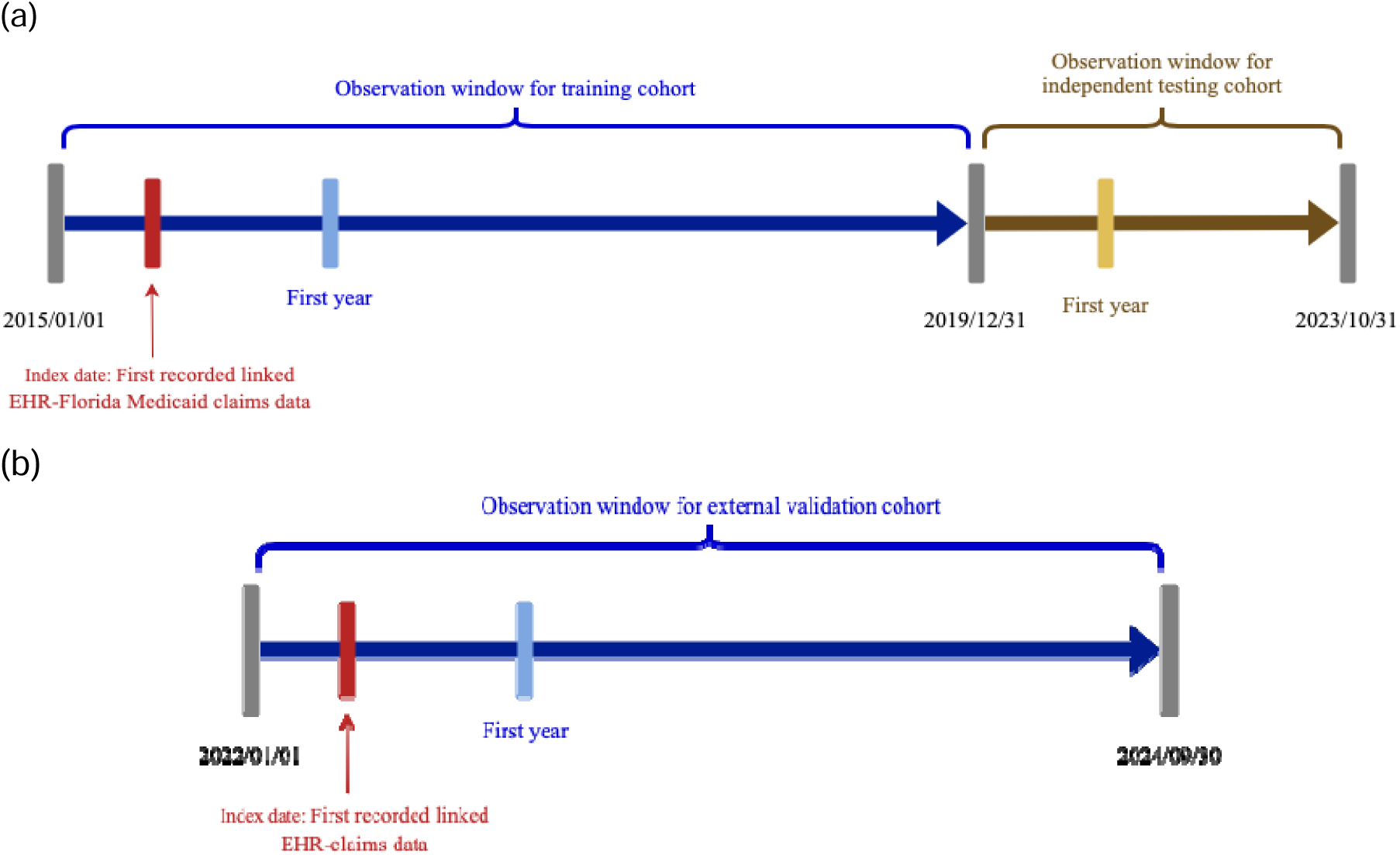
Overview of study design for (a) OneFlorida+ (b) REACHnet–LABlue.

### EHR data-continuity

To identify the optimal level of EHR data continuity, we applied the Harmonized Encounter Proportion Score (HEPS)^19^, a continuity metric developed to robustly estimate data completeness at the patient-level. HEPS is adapted from the Mean Proportion of Encounters Captured (MPEC)^8,10^ algorithm. MPEC calculates the proportion of healthcare encounters documented in the EHR compared to claims data:

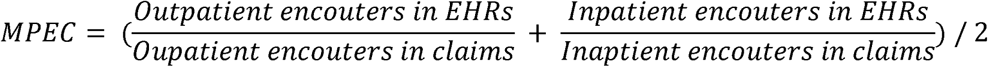

While MPEC is informative, it fails in cases where patients have no encounters in either the inpatient or outpatient setting during the observation period, leading to undefined (zero denominator) situations. HEPS has addressed this limitation by recursively handling such cases, ensuring a valid continuity score even when one type of encounter is absent.

The HEPS formula is defined as follows:

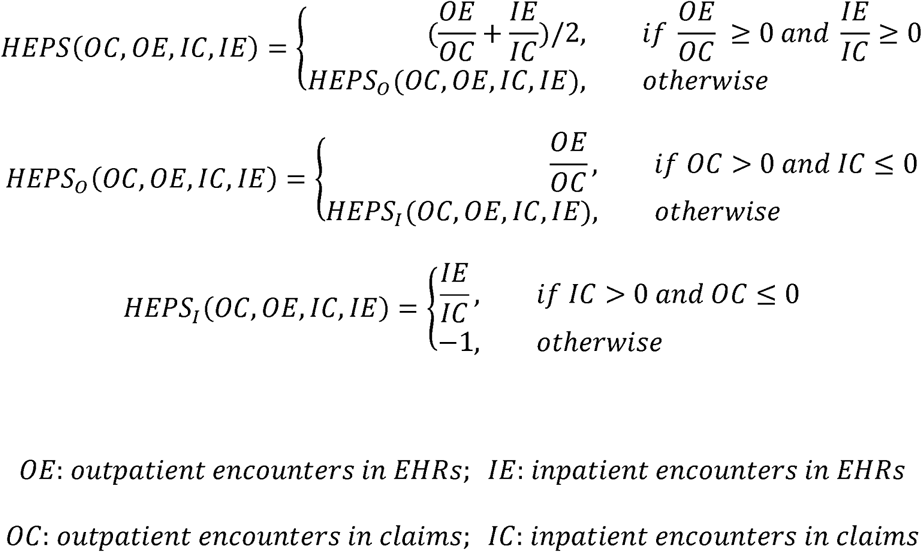

Using HEPS, we evaluated magnitude of EHR data continuity for each patient. We restricted the analytic cohort to patients with HEPS > 0% during the one-year observation window, because HEPS ≤ 0% indicates no captured inpatient and outpatient encounters in the EHR, the linked claims, or both. Such cases provide insufficient clinical information for continuity assessment and model development; therefore, they were excluded from the final analysis.

### Misclassification tests and evaluation metrics

To identify an optimal HEPS cutoff for defining high versus low EHR continuity, we adopted a methodology based on the framework proposed by Lin et al.^8,9,10^ We performed misclassification analyses using two types of clinical information commonly used in observational research:

(1) Comorbidity indices:
  a. Charlson Comorbidity Index (CCI) (range: 0 to 35),
  b. Elixhauser Comorbidity Index (ECI) (range: –19 to 89), and
  c. A combined score integrating both indices (range: –2 to 26), where higher scores are associated with increased mortality risk.^20^
(2) Clinical Variables: A set of 42 variables was assessed, comprising 30 chronic conditions^21^, and 12 frequently prescribed medications (see full list in Supplementary **Table S1**; definitions in Supplementary **Table S2**). These variables were evaluated during each patient’s first complete calendar year of data.

For each patient, comorbidity scores were calculated from both EHR-only data and linked EHR-claims data. The difference between the two (i.e., score_full **–** score_EHR) represents the extent to which comorbidity burden may be underestimated when using EHR data alone compared with using both claims and EHRs. We then compared the average differences across HEPS strata to evaluate the relationship between data continuity and score accuracy.

For the 42 selected clinical variables, we further assessed covariate balance between EHR-only data and linked EHR–claims data across HEPS deciles using two metrics: mean standardized differences (MSD) and sensitivity. The MSD compared classifications based on EHR-only data versus linked claims-EHR data; it quantified the difference between the two group means, standardized by their pooled standard deviation, and is commonly employed to evaluate covariate balance between comparison groups.^22^ A smaller MSD suggested a stronger agreement between data sources, indicating that the EHR alone provides sufficient information to approximate classifications derived from the more complete, linked dataset. Sensitivity of EHR coverage was defined as the proportion of patients identified with a given variable using EHR data alone, relative to those identified using the linked claims-EHR data:

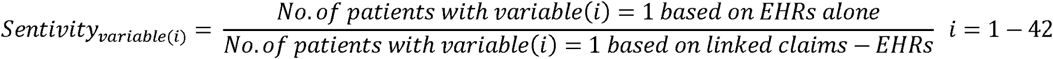

Given that the gold standard suggests classification based on all available data (both claims and EHR data), specificity was assumed to be 100%. However, incomplete coding in the EHR could result in reduced sensitivity, signaling potential misclassification.

Based on prior literature, a standardized difference < 0.10 is generally considered indicative of acceptable covariate balance and minimal confounding.^23,24^ A threshold of less than 0.10 for standardized differences across all variables was determined to indicate satisfactory balance of covariates in the context of achieving adequate confounding adjustment. Thus, we selected the HEPS threshold that achieved standardized differences below this benchmark across all 42 variables, defining this point as the optimal cutoff for high EHR data continuity.

### Model development, explainable AI and fairness optimization

To predict EHR data continuity, we identified a comprehensive set of candidate predictors based on clinical expertise^8^ and existing literature.^19^ Lin et al.^8^ proposed a list of candidate proxy indicators including (a) general examinations or routine care; (b) preventive interventions; (c) recording of diagnoses or medications in the EHR; (d) having a certain type and number of visits in the EHR; and (e) seeing the same provider repeatedly within the system (see Supplementary **Table S3** for detailed definitions). We further incorporated demographic, vital signs, clinical diagnoses variables from Huang et al.,^19^ including common diseases and provider-related factors. These predictors encompassed sociodemographic characteristics, vital signs, healthcare utilization patterns, and clinical indicators. The output variable was a binary indicator of high or low data continuity, as defined by the HEPS cutoff identified earlier.

To evaluate and optimize predictive performance and feasibility, we developed models using the predictor set identified from Lin et al., the predictor set identified from Huang et al., and the complete (combined) predictor set. Both logistic regression (LR) and XGBoost^25^ algorithms were used to develop prediction models. For each patient, the first complete calendar year of data following the index date (**Figure 1a**) was extracted, and the data were randomly split into training (70%) and validation (30%) cohorts. Model performance was assessed using standard classification metrics: area under the receiver operating characteristic curve (AUROC), F1 score, precision, and recall.

Model interpretability was enhanced using explainable AI techniques, specifically SHAP (SHapley Additive exPlanations)^26^ values, to identify key predictors influencing EHR continuity. Additionally, seven ML fairness metrics were applied to evaluate model fairness across racial and ethnic groups: non-Hispanic Black (NHB) vs non-Hispanic White (NHW), and Hispanic vs. NHW. The false-negative rate (FNR)—defined as the proportion of high-continuity patients incorrectly classified as having low continuity—was used as the primary measure of fairness, as false negatives result in the unnecessary exclusion of patients from data, potentially reducing cohort representativeness and introducing bias.

### External validation

Using the models developed from OneFlorida+ data, we assessed their generalizability in a linked EHR–claims dataset from the REACHnet–LABlue network. First, we replicated the misclassification analyses by (1) comparing mean differences in three comorbidity indices—CCI, ECI, and combined score—across HEPS strata to identify the minimal HEPS needed to achieve acceptable classification; and (2) evaluating covariate balance across 42 selected variables (30 chronic conditions and 12 commonly prescribed medications) using MSD, with <0.10 across all variables indicating satisfactory balance. Second, we evaluated transportability by applying the OneFlorida+-trained LR and XGBoost models to the REACHnet–LABlue predictors without recalibration. Model performance was evaluated across prespecified predictor sets (the predictor set identified from Lin et al.; the predictor set identified from Huang et al.) and the complete predictor set.

## RESULTS

A total of 5,462 OneFlorida+ adults with a complete first calendar year (2015–2019) and HEPS > 0% were included in the final analytic cohort; an additional 3,364 patients with a complete first calendar year (2020–2023) and HEPS > 0% comprised the testing cohort. In the REACHnet–LABlue system, 14,567 adults met eligibility criteria and were included in the external validation cohort. **Table 1** summarizes baseline characteristics. The OneFlorida+ training and testing cohorts were older than the REACHnet cohort (mean age 47.03 and 48.49 years, respectively, vs 45.14 years in REACHnet). The female proportion (∼65–67%) was similar across cohorts. Racial/ethnic composition differed: OneFlorida+ included larger Hispanic (25–26%) and NHB (29–31%) groups compared with the REACHnet cohort. Detailed provider-type and laboratory distributions are shown in **Supplementary Table S4**.

**Table 1.**
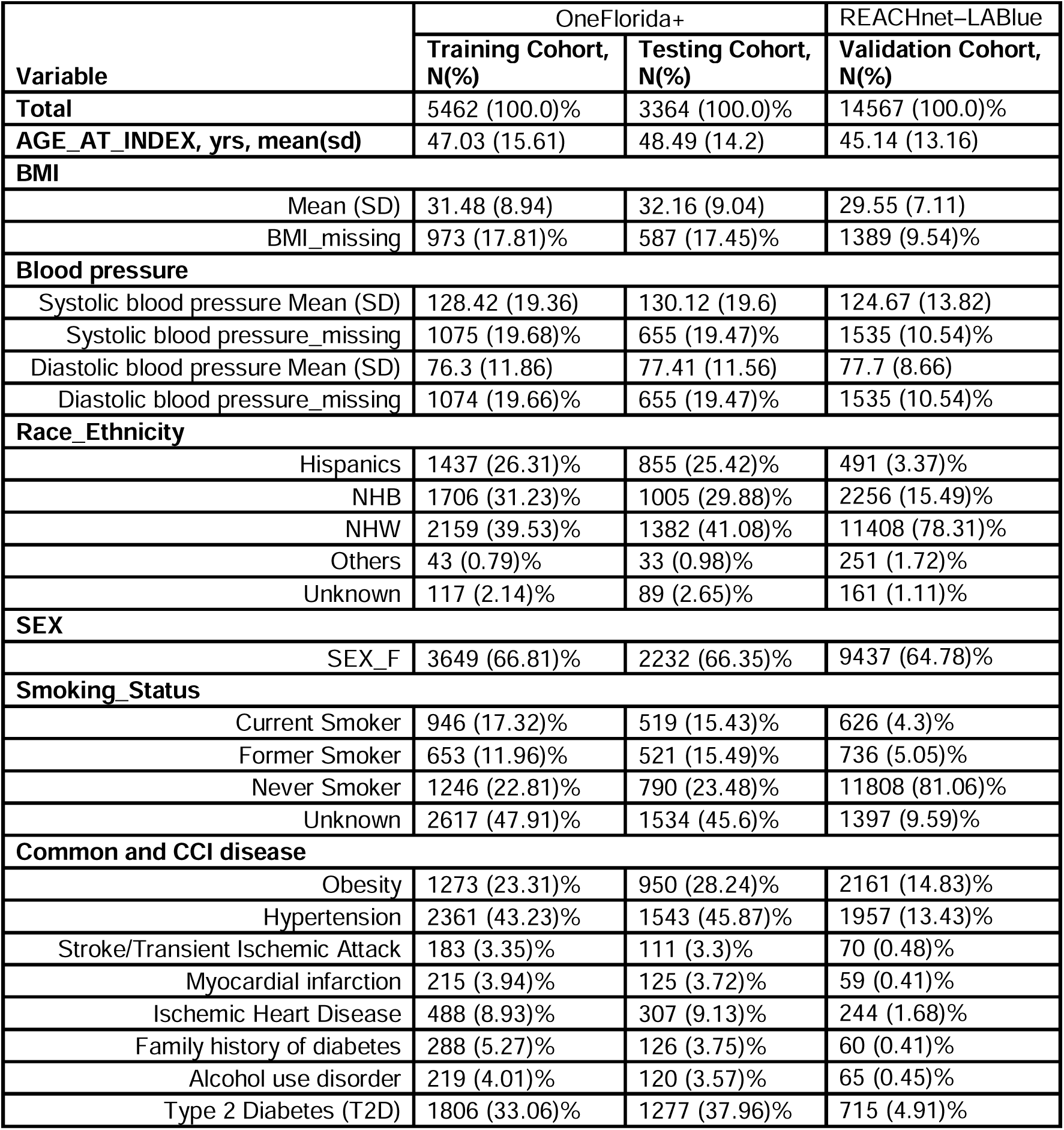
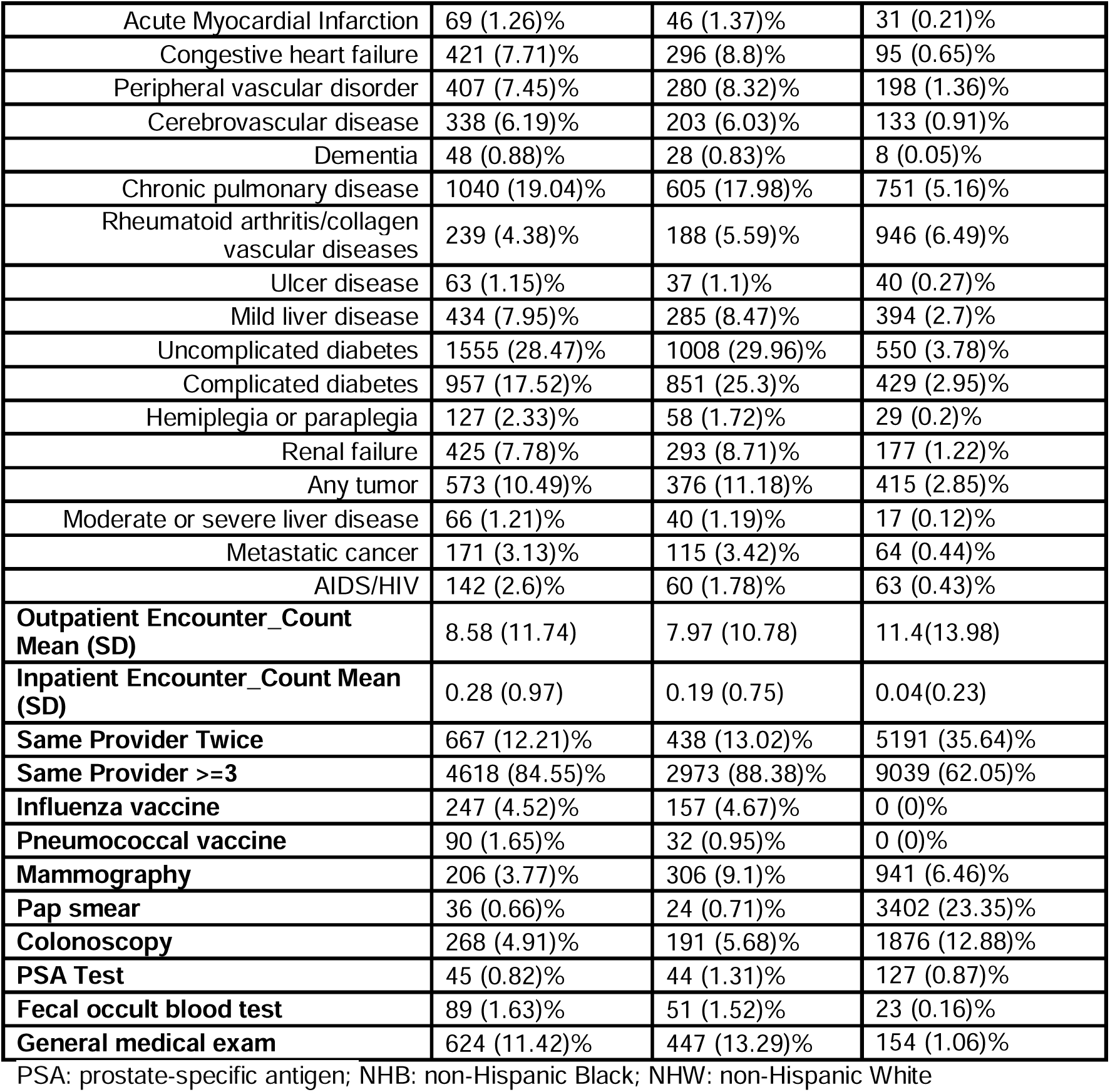
Population characteristics of training, testing, and validation cohort.

As shown in **Supplementary Figure S1a**, 91.9% of patients had HEPS scores below 50%, with a mean HEPS of 20%. In **Figure 2**, the mean differences in comorbidity scores between EHR-only data and linked EHR–claims data decreased as HEPS increased, stabilizing around a HEPS threshold of 30%. For patients with HEPS <10%, mean differences in CCI, ECI, and the combined score were 2.24, 3.14, and 1.46, which were 6.6-, 28.5-, and 9.7-fold higher, respectively, than those with HEPS ≥80% (0.34, 0.11, and 0.15). Full results with 95% confidence intervals across HEPS deciles are reported in **Supplementary Table S5**. A similar pattern was observed in the classification of the 42 selected clinical variables. **Figure 3** illustrates a declining trend in MSD comparing EHR-only with linked EHR–claims data as HEPS increased. Among patients with HEPS <10%, the average MSDs were 0.29 for comorbidity variables and 0.32 for medication variables–7.25-fold and 4-fold higher, respectively, than MSDs in patients with HEPS ≥80% (0.04 and 0.08). Full MSD results and confidence intervals by decile are provided in **Supplementary Table S6**. Using MSD of 0.10 as the cutoff,^23,24^ a minimum HEPS threshold of 30% was necessary to achieve acceptable classification of the selected variables in OneFlorida+, ensuring standardized differences were below the predefined cutoff for misclassification. Accordingly, we defined patients with HEPS ≥30% as the “EHR high-continuity cohort.” Sensitivity analysis, presented in **Supplementary Figure S2**, further confirmed this pattern, showing a consistent increase in the average sensitivity of EHRs in capturing codes for the 42 selected variables as HEPS increased, validating HEPS as a strong proxy for data completeness. Lastly, we examined the distribution of data continuity across racial and ethnic groups (**Table 2**). The NHW subgroup had a higher proportion of patients with high EHR continuity compared with the NHB and other racial/ethnic groups. Both NHW and NHB patients were more frequently represented in the high-continuity group than in the low-continuity group, although the disparity remained more pronounced for NHW.

**Figure 2.**
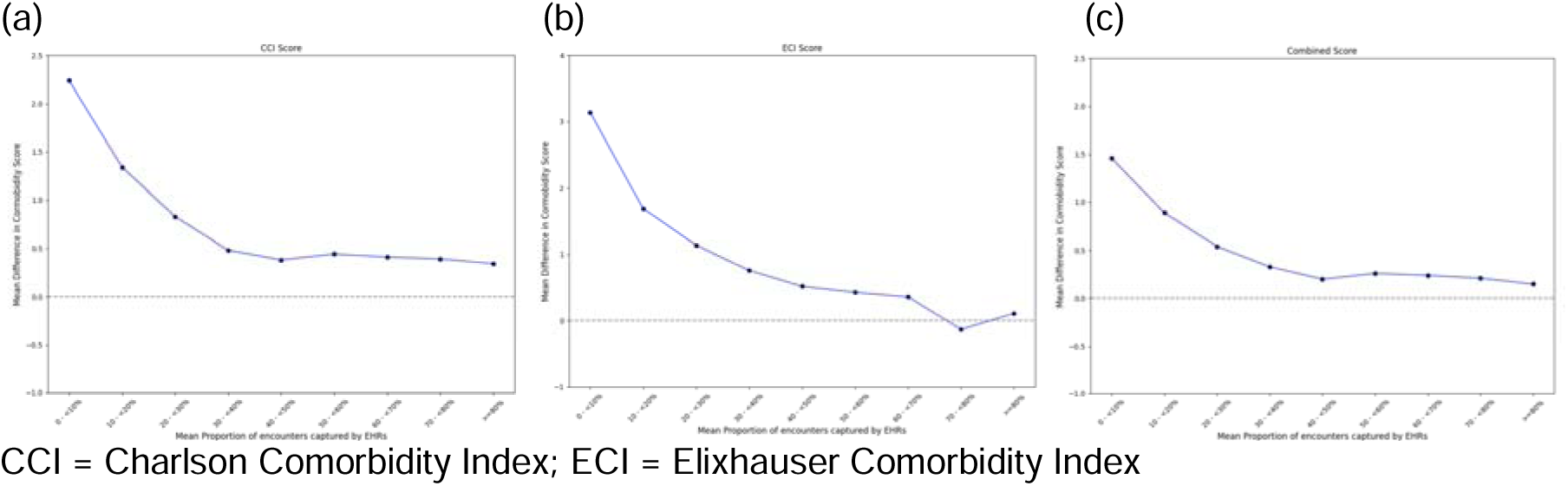
Mean difference by HEPS score deciles in (a) CCI score, (b) ECI score, (c) Combined score in OneFlorida+.

**Figure 3.**
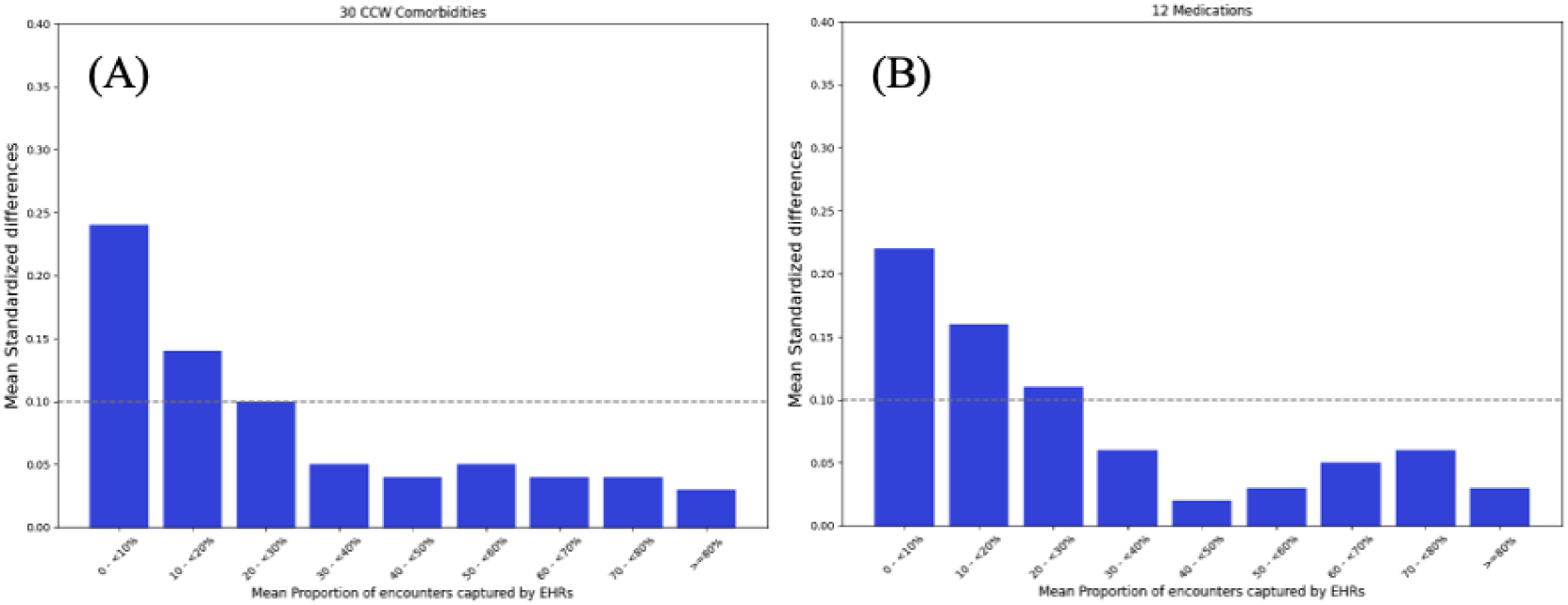
Mean standardized difference between EHR-claims linked vs EHR-only data for (A) comorbidities and (B)medications in OneFlorida+.

**Table 2.**
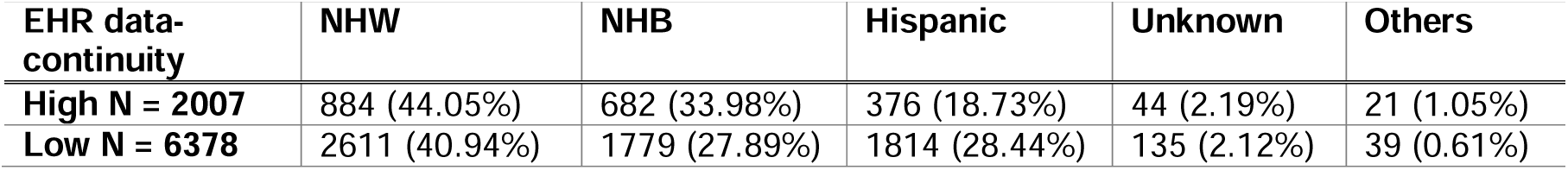
Number of patients and their distribution by high versus low data-continuity in racial subgroups in OneFlorida+.

| EHR data-continuity | NHW | NHB | Hispanic | Unknown | Others |
| --- | --- | --- | --- | --- | --- |
| High N = 2007 | 884 (44.05%) | 682 (33.98%) | 376 (18.73%) | 44 (2.19%) | 21 (1.05%) |
| Low N = 6378 | 2611 (40.94%) | 1779 (27.89%) | 1814 (28.44%) | 135 (2.12%) | 39 (0.61%) |

While both LR and XGBoost continuity-prediction algorithms demonstrated excellent performance in identifying patients with high EHR continuity in both internal and external validation (**Table 3**), XGBoost consistently outperformed LR across all predictor sets. Models trained with the complete predictor set achieved the best performance, with improved AUROC and F1-score compared with models trained on the predictor set identified from Lin et al.’s paper^8^ or on the predictor set identified from Huang et al.’s paper.^19^ Overall, the XGBoost model using combined data showed the best predictive performance, achieving an AUROC of 0.77.

**Table 3.** Comparative performance of the models in OneFlorida+.

| Data | Model | AUROC | F1-Score | Specificity | Accuracy |
| --- | --- | --- | --- | --- | --- |
| **Complete predictor set** | XGBoost | 0.77 | 0.54 | 0.72 | 0.71 |
|  | LR | 0.75 | 0.53 | 0.68 | 0.68 |
| **Predictor set (Huang et al.)** | XGBoost | 0.77 | 0.53 | 0.73 | 0.71 |
|  | LR | 0.73 | 0.51 | 0.67 | 0.67 |
| **Predictor set (Lin et al.)** | XGBoost | 0.73 | 0.51 | 0.72 | 0.69 |
|  | LR | 0.72 | 0.51 | 0.69 | 0.68 |

AUROC: area under the receiver operating characteristic curve; LR: logistic regression

Subsequently, using the XGBoost model with the complete predictor set, we assessed model interpretability with SHAP values (**Figure 4**) to identify key predictors of high EHR continuity. Significant predictors included features from both data sources, such as “outpatient encounter count”, “triglycerides”, and “having BMI records”. These findings highlight the importance of incorporating clinical and encounter-related variables in predictive modeling for EHR continuity. SHAP analyses for LR and additional datasets are provided in **Supplementary Figure S3 to S7**. We further evaluated model fairness by examining the FNR across racial and ethnic subgroups in the XGBoost models (**Table 4**), with additional metrics available in **Supplementary Table S7**. Overall, the algorithm appeared biased against the Hispanic group relative to NHW; fairness metrics improved significantly after applying the adversarial debiasing technique.

**Figure 4.**
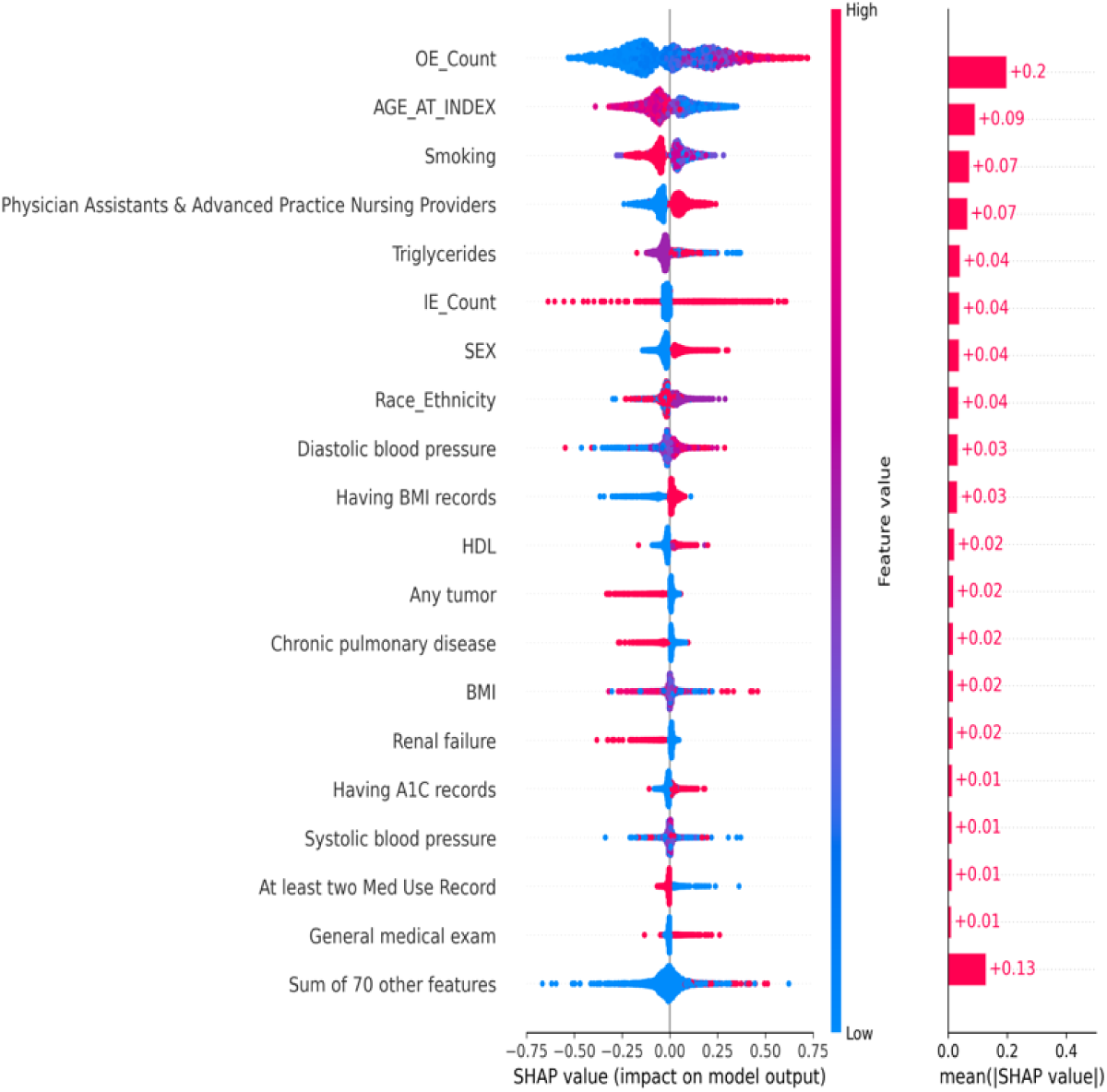
SHAP values for XGBoost on the complete predictor set.

**Table 4.**
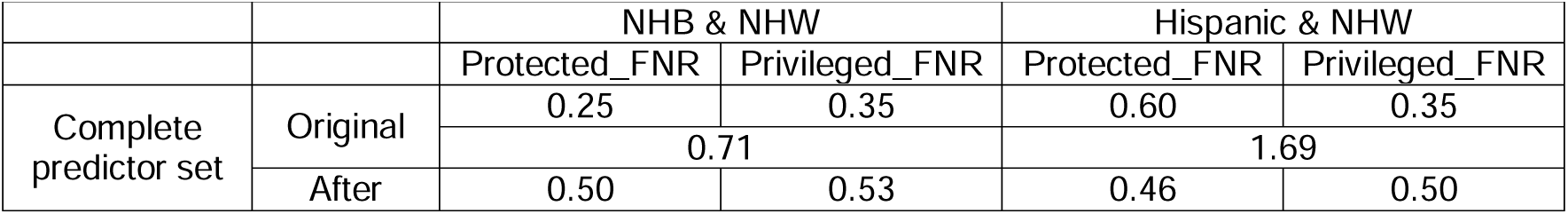
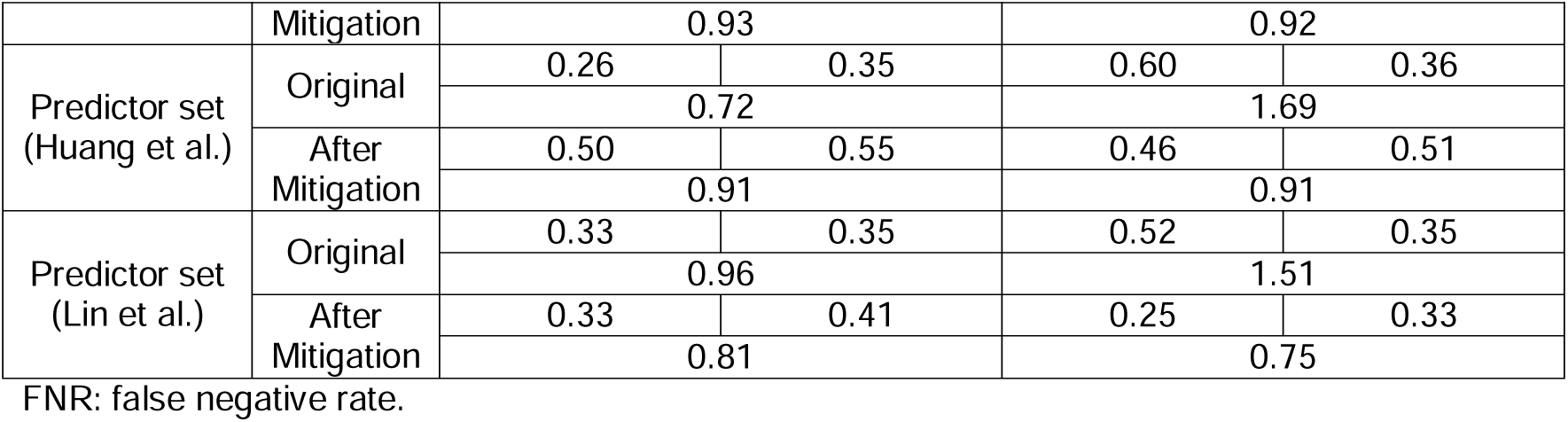
FNR across race-ethnicity groups in XGBoost models.

In the external validation using REACHnet data, 77.9% of patients had HEPS <50% (mean 31%; **Supplementary Figure S1b**). Mean differences between EHR-only and linked EHR–claims comorbidity scores declined with increasing HEPS and stabilized at around 30% (**Supplementary Figure S8**); a consistent pattern was observed for the 42 variables, with MSD decreasing as HEPS increased (**Supplementary Figure S9**). Because MSDs for the 12 medication variables were uniformly small (<0.05), the 30 condition variables informed threshold selection; applying the prespecified MSD <0.10 criterion, HEPS ≥30% was required for acceptable classification, defining the “EHR high-continuity cohort.” By race/ethnicity groups (**Supplementary Table S8**), the Hispanic subgroup had the highest high-continuity proportion, whereas NHB and other groups had <20%. Model performance was stable across predictor sets (**Supplementary Table S9**): the predictor set identified from Huang et al. showed the strongest transportability (XGBoost AUROC 0.81, F1 0.75; LR AUROC 0.78, F1 0.73), the complete predictor set offered similar discrimination (AUROC 0.74–0.76) with XGBoost maintaining higher specificity and accuracy than LR (0.71/0.68 vs 0.41/0.65), and the predictor set identified from Lin et al. yielded lower performance (LR AUROC 0.76; XGBoost AUROC 0.65 with higher specificity 0.76).

## DISCUSSION

Using linked EHR–claims data, we quantified misclassification across comorbidity indices and 42 clinical variables and identified a pragmatic threshold (HEPS ≥30%) defining an “EHR high-continuity cohort.” We then developed person-level continuity classifiers that achieved excellent discrimination in internal testing (best AUROC ≈0.77) and transported well to an independent network.

In this study, we evaluated the magnitude of completeness of patient medical information captured in the OneFlorida+ EHR system by estimating EHR data continuity using a novel HEPS approach – a data continuity–measuring index adapted from prior MPEC algorithm.^8,10^ Our results showed that the mean HEPS in the study cohort was 20%, with the 91.9% of patients having HEPS values below 50%. These findings were consistent with previous studies reporting mean MPEC of 27% and 26% in Massachusetts and North Carolina EHR systems, with 74% and 78% of patients having MPEC below 50%.^10^ Taken together, these estimates suggest a single network does usually not comprehensively capture all clinically relevant encounters for most patients in the cohort. Despite the relatively low HEPS distribution, we found that a HEPS threshold of 30%, was sufficient to achieve acceptable low misclassification (MSD < 0.10) for 42 commonly used comorbidity and medication variables. This threshold was further confirmed by trends observed in sensitivity analyses and mean difference analysis of comorbidity score. Consistent patterns were observed in external validation using the REACHnet–LABlue dataset, where the mean HEPS was 30% and misclassification decreased monotonically with increasing HEPS. These findings suggest that HEPS offer a practical approach for researchers to quantify the extent of information bias associated with low EHR continuity and to assess the potential impact of data incompleteness on study validity.

Using data from the OneFlorida+ EHR system, we developed and validated machine learning models to predict high EHR data continuity. Models trained on the complete predictor set performed slightly better than those trained on either the predictor set identified from Lin et al. or on the predictor set identified from Huang et al.. While all models achieved comparable predictive performance, the predictor set identified from Lin et al. yielded the lowest accuracy, and the predictor set identified from Huang et al. demonstrated slightly greater predictive power. These findings suggest that both individual data domains contribute meaningfully to model performance, and their combination provides added value in improving prediction accuracy. External validation in REACHnet–LABlue showed that these models retained stable discrimination, with the predictor set identified from Huang et al. exhibiting the strongest transportability (e.g., XGBoost AUROC 0.81), and the complete predictor set yielding similar discrimination with more balanced operating characteristics for XGBoost than for LR. Overall, our results demonstrate the feasibility of using EHR-only data to detect high EHR continuity, offering a scalable approach to improve cohort selection and reduce information bias in real-world data research.

Beyond research validity, continuity prediction models have potential operational and clinical applications. Health systems could use HEPS or similar measures to identify patients whose fragmented care may lead to gaps in chronic disease management, medication reconciliation, or preventive screening. Integrating continuity scores into population health dashboards or risk stratification tools could help care managers proactively engage patients likely to have incomplete EHR documentation or inconsistent follow-up. From a research perspective, embedding continuity-aware cohort selection into real-world evidence pipelines may enhance both study rigor and fairness by ensuring equitable representation of patients with reliable data capture.

This study has several limitations that inform next steps. First, because HEPS relies on EHR and claims linkage, it may miss out-of-network care. In addition, we developed our EHR continuity prediction model using EHR–Medicaid-claims–linked data and externally validated it in EHR–commercial-claims–linked data. However, it is unclear whether the model can be generalized to older populations enrolled in Medicare.

## CONCLUSION

In conclusion, we developed a fair, generalizable ML model that exhibits excellent predictive performance in identifying patients with high EHR continuity. These findings underscore the potential of our model to enhance the quality of EHR data for research by accurately identifying individuals with sufficient and reliable clinical documentation.

## Data Availability

Data set Available through OneFlorida+Clinical Research Network (email) and Research Action of Health Network (link, https://www.reachnet.org/resources/forms)

## ACKNOWLEDGMENT

None

## AUTHOR CONTRIBUTIONS

Conceptualization, J. Guo, J. Bian, Y. Huang, and L. Shi; methodology, Y. Huang and J. Guo; formal analysis, YA. Lee and T. Tang; data curation, YA. Lee and T. Tang; resources, J. Guo and L. Shi; writing – initial draft, YA. Lee and T. Tang; critical review and editing, J. Guo and L. Shi; supervision: J. Guo and L. Shi. All authors have read and agreed to the published version of the manuscript.

