## Supplemental Materials for "Develop and Validate A Fair Machine Learning Model to Indentify Patients with High Care-Continuity in Electronic Health Records Data"

1. **Table S1**: Selected variables for misclassification test
2. **Table S2**: Definition of 42 selected variables
3. **Table S3**: Definitions of the candidate predictors for EHR care-continuity
4. **Table S4**: Population characteristics of training, testing, and validation cohort
5. **Table S5**: Mean difference and 95% confidence interval by each HEPS decile in CCI score, ECI score, and Combined score in OneFlorida+
6. **Table S6**: Mean standardized difference and 95% confidence interval by each HEPS decile in 42 selected variables in OneFlorida+
7. **Table S7**: Fairness assessment across race-ethnicity groups in XGBoost models
8. **Table S8**: Number of patients and their distribution by high versus low data-continuity in racial subgroups in REACHnet–LABlue
9. **Table S9**: Comparative performance of the models in REACHnet–LABlue.
10. **Figure S1**: Number of patients by HEPS score category in the (a) OneFlorida+ cohort, (b) REACHnet–LABlue cohort
11. **Figure S2**: Sensitivity of capturing clinical information in relation of EHR-continuity in OneFlorida+
12. **Figure S3**: SHAP values for XGBoost on the predictor set identified from Huang et al.
13. **Figure S4**: SHAP values for XGBoost on the predictor set identified from Lin et al.
14. **Figure S5**: SHAP values for Logistic Regression on the complete predictor set.
15. **Figure S6**: SHAP values for Logistic Regression on the predictor set identified from Huang et al.
16. **Figure S7**: SHAP values for Logistic Regression on the predictor set identified from Lin et al.
17. **Figure S8**: Mean difference by HEPS score deciles in (a) CCI score, (b) ECI score, (c) Combined score in REACHnet–LABlue
18. **Figure S9**: Mean standardized difference between EHR-claims linked vs EHR-only data for (A) comorbidities and (B) medications in REACHnet–LABlue.

**Table S1.** **Selected variables for misclassification test**

| 30 comorbidity variables | CCW Chronic Conditions:  Acute Myocardial Infarction, Alzheimer's Disease, Anemia, Asthma, Atrial Fibrillation and Flutter, Benign Prostatic Hyperplasia, Breast Cancer, Colorectal Cancer, Endometrial Cancer, Lung Cancer, Prostate Cancer, Urologic (Kidney, Renal Pelvis, and Ureter) Cancer, Cataract, Chronic Kidney Disease, Chronic Obstructive Pulmonary Disease, Depression, Bipolar (or Other Depressive Mood Disorders), Diabetes, Glaucoma, Heart Failure and Non-Ischemic Heart Disease, Hip/Pelvic Fracture, Hyperlipidemia, Hypertension, Hypothyroidism, Ischemic Heart Disease, Non-Alzheimer's Dementia, Osteoporosis With or Without Pathological Fracture, Parkinson's Disease and Secondary Parkinsonism , Pneumonia (All-cause), Rheumatoid Arthritis/Osteoarthritis, Stroke/Transient Ischemic Attack |
| --- | --- |
| 12 medication variables | Antiplatelet agents, Oral anticoagulants, Antihypertensives, statins, NSAIDs, Opioids, Antidepressants, Antipsychotics, Anti-dementia, PPIs, GLDs, Antidiabetics |

NSAIDs = nonsteroidal anti-inflammatory drugs; PPIs = Proton pump inhibitors; GLDs = Glucose lowering drugs

**Table S2. Definition of 42 selected variables**

| **CCW Chronic condition variables** | **Definition** |
| --- | --- |
| Acute Myocardial Infarction | I21.01, I21.02, I21.09, I21.11, I21.19, I21.21, I21.29, I21.3, I21.4, I21.9, I21.A1, I21.A9, I21.B, I22.0, I22.1, I22.2, I22.8, I22.9, I23.0, I23.1, I23.2, I23.3, I23.4, I23.5, I23.6, I23.7, I23.8 (any DX on the claim) |
| Alzheimer's Disease | G30.0, G30.1, G30.8, G30.9 (any DX on the claim) |
| Anemia | C94.6, D46.0, D46.1, D46.20, D46.21, D46.22, D46.4, D46.9, D46.A, D46.B, D46.C, D46.Z, D47.4, D50.0, D50.1, D50.8, D50.9, D51.0, D51.1, D51.2, D51.3, D51.8, D51.9, D52.0, D52.1, D52.8, D52.9, D53.0, D53.1, D53.2, D53.8, D53.9, D55.0, D55.1, D55.2, D55.21, D55.29, D55.3, D55.8, D55.9, D56.0, D56.1, D56.2, D56.3, D56.4, D56.5, D56.8, D56.9, D57.00, D57.01, D57.02, D57.03, D57.04, D57.09, D57.1, D57.20, D57.211, D57.212, D57.213, D57.214, D57.218, D57.219, D57.3, D57.40, D57.411, D57.412, D57.413, D57.414, D57.418, D57.419, D57.42, D57.431, D57.432, D57.433, D57.434, D57.438, D57.439, D57.44, D57.451, D57.452, D57.453, D57.454, D57.458, D57.459, D57.80, D57.811, D57.812, D57.813, D57.814, D57.818, D57.819, D58.0, D58.1, D58.2, D58.8, D58.9, D59.0, D59.1, D59.10, D59.11, D59.12, D59.13, D59.19, D59.2, D59.3, D59.30, D59.31, D59.32, D59.39, D59.4, D59.5, D59.6, D59.8, D59.9, D60.0, D60.1, D60.8, D60.9, D61.01, D61.02, D61.09, D61.1, D61.2, D61.3, D61.810, D61.811, D61.818, D61.82, D61.89, D61.9, D63.0, D63.1, D63.8, D64.0, D64.1, D64.2, D64.3, D64.4, D64.81, D64.89, D64.9, D75.81 (any DX on the claim) |
| Asthma | J45.20, J45.21, J45.22, J45.30, J45.31, J45.32, J45.40, J45.41, J45.42, J45.50, J45.51, J45.52, J45.901, J45.902, J45.909, J45.990, J45.991, J45.998 (any DX on the claim) |
| Atrial Fibrillation and Flutter | I48.0, I48.1, I48.11, I48.19, I48.2, I48.20, I48.21, I48.3, I48.4, I48.91 (any DX on the claim) |
| Benign Prostatic Hyperplasia | N40.0, N40.1, N40.2, N40.3 (any DX on the claim)  EXCLUSION: If any of the qualifying claims also have an ICD-10 diagnosis of D29.1, then EXCLUDE |
| Cancer, Breast | C50.011, C50.012, C50.019, C50.021, C50.022, C50.029, C50.111, C50.112, C50.119, C50.121, C50.122, C50.129, C50.211, C50.212, C50.219, C50.221, C50.222, C50.229, C50.311, C50.312, C50.319, C50.321, C50.322, C50.329, C50.411, C50.412, C50.419, C50.421, C50.422, C50.429, C50.511, C50.512, C50.519, C50.521, C50.522, C50.529, C50.611, C50.612, C50.619, C50.621, C50.622, C50.629, C50.811, C50.812, C50.819, C50.821, C50.822, C50.829, C50.911, C50.912, C50.919, C50.921, C50.922, C50.929, D05.00, D05.01, D05.02, D05.10, D05.11, D05.12, D05.80, D05.81, D05.82, D05.90, D05.91, D05.92, Z17.0,  Z17.1, Z19.1, Z19.2, Z85.3, Z86.000 (any DX on the claim) |
| Cancer, Colorectal | C18.0, C18.1, C18.2, C18.3, C18.4, C18.5, C18.6, C18.7, C18.8, C18.9, C19, C20, C49.A4, C49.A5, D01.0, D01.1, D01.2, Z85.030, Z85.038, Z85.040, Z85.048 (any DX on the claim) |
| Cancer, Endometrial | C54.0, C54.1, C54.2, C54.3, C54.8, C54.9, D07.0, Z85.42 (any DX on the claim) |
| Cancer, Lung | C34.00, C34.01, C34.02, C34.10, C34.11, C34.12, C34.2, C34.30, C34.31, C34.32, C34.80, C34.81, C34.82, C34.90, C34.91, C34.92, D02.20, D02.21, D02.22, Z85.110, Z85.118 (any DX on the claim) |
| Cancer, Prostate | C61, D07.5, Z85.46 (any DX on the claim) |
| Cancer, Urologic (Kidney, Renal Pelvis, and Ureter) | C64.1, C64.2, C64.9, C65.1, C65.2, C65.9, C66.1, C66.2, C66.9, C68.8, C68.9, D09.10, D09.19, Z85.520, Z85.528, Z85.53, Z85.54,  Z85.59 (any DX on the claim) |
| Cataract | E08.36, E09.36, E10.36, E11.36, E13.36, H25.011, H25.012, H25.013, H25.019, H25.031, H25.032, H25.033, H25.039, H25.041, H25.042, H25.043, H25.049, H25.091, H25.092, H25.093, H25.099, H25.10, H25.11, H25.12, H25.13, H25.20, H25.21, H25.22, H25.23, H25.811, H25.812, H25.813, H25.819, H25.89, H25.9, H26.001, H26.002, H26.003, H26.009, H26.011, H26.012, H26.013, H26.019, H26.031, H26.032, H26.033, H26.039, H26.041, H26.042, H26.043, H26.049, H26.051, H26.052, H26.053, H26.059, H26.061, H26.062, H26.063, H26.069, H26.09, H26.101, H26.102, H26.103, H26.109, H26.111, H26.112, H26.113,  H26.119, H26.121, H26.122, H26.123, H26.129, H26.131, H26.132, H26.133, H26.139, H26.20, H26.211, H26.212, H26.213, H26.219, H26.221, H26.222, H26.223, H26.229, H26.30, H26.31, H26.32, H26.33, H26.40, H26.411, H26.412, H26.413, H26.419, H26.491, H26.492, H26.493, H26.499, H26.8, H26.9, Q12.0 (any DX on the claim) |
| Chronic Kidney Disease | A18.11, A52.75, B52.0, E08.21, E08.22, E08.29, E09.21, E09.22, E09.29, E10.21, E10.22, E10.29, E11.21, E11.22, E11.29, E13.21, E13.22, E13.29, I12.0, I12.9, I13.0, I13.10, I13.11, I13.2, K76.7, M10.30, M10.311, M10.312, M10.319, M10.321, M10.322, M10.329, M10.331, M10.332, M10.339, M10.341, M10.342, M10.349, M10.351, M10.352, M10.359, M10.361, M10.362, M10.369, M10.371, M10.372, M10.379, M10.38, M10.39, M32.14, M32.15, M35.04, M35.0A, N01.0, N01.1, N01.2, N01.3, N01.4, N01.5, N01.6, N01.7, N01.8, N01.9, N01.A, N02.0, N02.1, N02.2, N02.3, N02.4, N02.5, N02.6, N02.7, N02.8, N02.9, N02.A, N02.B1, N02.B2, N02.B3, N02.B4, N02.B5, N02.B6, N02.B9, N03.0, N03.1, N03.2, N03.3, N03.4, N03.5, N03.6, N03.7, N03.8, N03.9, N03.A, N04.0, N04.1, N04.2, N04.20, N04.21, N04.22, N04.29, N04.3, N04.4, N04.5, N04.6, N04.7, N04.8, N04.9, N04.A, N05.0, N05.1, N05.2, N05.3, N05.4, N05.5, N05.6, N05.7, N05.8, N05.9, N05.A, N06.0, N06.1, N06.2, N06.20, N06.21, N06.22, N06.29, N06.3, N06.4, N06.5, N06.6, N06.7, N06.8, N06.9, N06.A, N07.0, N07.1, N07.2, N07.3, N07.4, N07.5, N07.6, N07.7, N07.8, N07.9, N07.A, N08, N14.0, N14.1, N14.11, N14.19, N14.2, N14.3, N14.4, N15.0, N15.8, N15.9, N16, N18.1, N18.2, N18.3, N18.30, N18.31, N18.32, N18.4, N18.5, N18.6, N18.9, N25.1, N25.89, N25.9, N26.1, N26.9, N99.0, Q61.02, Q61.11, Q61.19, Q61.2, Q61.3, Q61.4, Q61.5, Q61.8 (any DX on the claim) |
| Chronic Obstructive Pulmonary Disease | J40, J41.0, J41.1, J41.8, J42, J43.0, J43.1, J43.2, J43.9, J44.0, J44.1, J44.81, J44.89, J44.9, J47.0, J47.1, J47.9, J98.2, J98.3 (any DX don the claim) |
| Depression, Bipolar, or Other Depressive Mood Disorders | F06.31, F06.32, F31.0, F31.10, F31.11, F31.12, F31.13, F31.2, F31.30, F31.31, F31.32, F31.4, F31.5, F31.60, F31.61, F31.62, F31.63, F31.64, F31.71, F31.73, F31.75, F31.76, F31.77, F31.78, F31.81, F31.89, F31.9, F32.0, F32.1, F32.2, F32.3, F32.4, F32.5, F32.8, F32.89, F32.9, F32.A, F33.0, F33.1, F33.2, F33.3, F33.40, F33.41, F33.42, F33.8, F33.9, F34.0, F34.1, F43.21, F43.23 (any DX on the claim) |
| Diabetes | E08.00, E08.01, E08.10, E08.11, E08.21, E08.22, E08.29, E08.311, E08.319, E08.321, E08.3211, E08.3212, E08.3213, E08.3219, E08.329, E08.3291, E08.3292, E08.3293, E08.3299, E08.331, E08.3311, E08.3312, E08.3313, E08.3319, E08.339, E08.3391,  E08.3392, E08.3393, E08.3399, E08.341, E08.3411, E08.3412, E08.3413, E08.3419, E08.349, E08.3491, E08.3492, E08.3493, E08.3499, E08.351, E08.3511, E08.3512, E08.3513, E08.3519, E08.3521, E08.3522, E08.3523, E08.3529, E08.3531, E08.3532,  E08.3533, E08.3539, E08.3541, E08.3542, E08.3543, E08.3549, E08.3551, E08.3552, E08.3553, E08.3559, E08.359, E08.3591, E08.3592, E08.3593, E08.3599, E08.36, E08.37X1, E08.37X2, E08.37X3, E08.37X9, E08.39, E08.40, E08.41, E08.42, E08.43,  E08.44, E08.49, E08.51, E08.52, E08.59, E08.610, E08.618, E08.620, E08.621, E08.622, E08.628, E08.630, E08.638, E08.641, E08.649, E08.65, E08.69, E08.8, E08.9, E09.00, E09.01, E09.10, E09.11, E09.21, E09.22, E09.29, E09.311, E09.319, E09.321, E09.3211, E09.3212, E09.3213, E09.3219, E09.329, E09.3291, E09.3292, E09.3293, E09.3299, E09.331, E09.3311, E09.3312, E09.3313, E09.3319, E09.339, E09.3391, E09.3392, E09.3393, E09.3399, E09.341, E09.3411, E09.3412, E09.3413, E09.3419, E09.349, E09.3491, E09.3492, E09.3493, E09.3499, E09.351, E09.3511, E09.3512, E09.3513, E09.3519, E09.3521, E09.3522, E09.3523, E09.3529, E09.3531, E09.3532, E09.3533, E09.3539, E09.3541, E09.3542, E09.3543, E09.3549, E09.3551, E09.3552, E09.3553, E09.3559, E09.359, E09.3591, E09.3592, E09.3593, E09.3599, E09.36, E09.37X1, E09.37X2, E09.37X3, E09.37X9, E09.39, E09.40, E09.41, E09.42, E09.43, E09.44, E09.49, E09.51, E09.52, E09.59, E09.610, E09.618, E09.620, E09.621, E09.622, E09.628, E09.630, E09.638, E09.641, E09.649, E09.65, E09.69, E09.8, E09.9, E10.10, E10.11, E10.21, E10.22, E10.29, E10.311, E10.319, E10.321, E10.3211, E10.3212, E10.3213, E10.3219, E10.329, E10.3291, E10.3292, E10.3293, E10.3299, E10.331, E10.3311, E10.3312, E10.3313, E10.3319, E10.339, E10.3391, E10.3392, E10.3393, E10.3399, E10.341, E10.3411, E10.3412, E10.3413, E10.3419, E10.349, E10.3491, E10.3492, E10.3493, E10.3499, E10.351, E10.3511, E10.3512, E10.3513, E10.3519, E10.3521, E10.3522, E10.3523, E10.3529, E10.3531, E10.3532, E10.3533, E10.3539, E10.3541, E10.3542, E10.3543, E10.3549, E10.3551, E10.3552, E10.3553, E10.3559, E10.359, E10.3591, E10.3592, E10.3593, E10.3599, E10.36, E10.37X1, E10.37X2, E10.37X3, E10.37X9, E10.39, E10.40, E10.41, E10.42, E10.43, E10.44, E10.49, E10.51, E10.52, E10.59, E10.610, E10.618, E10.620, E10.621, E10.622, E10.628, E10.630, E10.638, E10.641, E10.649, E10.65, E10.69, E10.8, E10.9, E11.00, E11.01, E11.10, E11.11, E11.21, E11.22, E11.29, E11.311, E11.319, E11.321, E11.3211, E11.3212, E11.3213, E11.3219, E11.329, E11.3291, E11.3292, E11.3293, E11.3299, E11.331, E11.3311, E11.3312, E11.3313, E11.3319, E11.339, E11.3391, E11.3392, E11.3393, E11.3399, E11.341, E11.3411, E11.3412, E11.3413, E11.3419, E11.349, E11.3491, E11.3492, E11.3493, E11.3499, E11.351, E11.3511, E11.3512, E11.3513, E11.3519, E11.3521, E11.3522, E11.3523, E11.3529, E11.3531, E11.3532, E11.3533, E11.3539, E11.3541, E11.3542, E11.3543, E11.3549, E11.3551, E11.3552, E11.3553, E11.3559, E11.359, E11.3591, E11.3592, E11.3593, E11.3599, E11.36, E11.37X1, E11.37X2, E11.37X3, E11.37X9, E11.39, E11.40, E11.41, E11.42, E11.43, E11.44, E11.49, E11.51, E11.52, E11.59, E11.610, E11.618, E11.620, E11.621, E11.622, E11.628, E11.630, E11.638, E11.641, E11.649, E11.65, E11.69,  E11.8, E11.9, E13.00, E13.01, E13.10, E13.11, E13.21, E13.22, E13.29, E13.311, E13.319, E13.321, E13.3211, E13.3212, E13.3213, E13.3219, E13.329, E13.3291, E13.3292, E13.3293, E13.3299, E13.331, E13.3311, E13.3312, E13.3313, E13.3319,  E13.339, E13.3391, E13.3392, E13.3393, E13.3399, E13.341, E13.3411, E13.3412, E13.3413, E13.3419, E13.349, E13.3491, E13.3492, E13.3493, E13.3499, E13.351, E13.3511, E13.3512, E13.3513, E13.3519, E13.3521, E13.3522, E13.3523, E13.3529,  E13.3531, E13.3532, E13.3533, E13.3539, E13.3541, E13.3542, E13.3543, E13.3549, E13.3551, E13.3552, E13.3553, E13.3559, E13.359, E13.3591, E13.3592, E13.3593, E13.3599, E13.36, E13.39, E13.40, E13.41, E13.42, E13.43, E13.44, E13.49, E13.51,  E13.52, E13.59, E13.610, E13.618, E13.620, E13.621, E13.622, E13.628, E13.630, E13.638, E13.641, E13.649, E13.65, E13.69, E13.8, E13.9 (any DX on the claim) |
| Glaucoma | H40.011, H40.012, H40.013, H40.019, H40.021, H40.022, H40.023, H40.029, H40.041, H40.042, H40.043, H40.049, H40.051, H40.052, H40.053, H40.059, H40.10X0, H40.10X1, H40.10X2, H40.10X3, H40.10X4, H40.1110, H40.1111, H40.1112, H40.1113, H40.1114, H40.1120, H40.1121, H40.1122, H40.1123, H40.1124, H40.1130, H40.1131, H40.1132, H40.1133, H40.1134,  H40.1190, H40.1191, H40.1192, H40.1193, H40.1194, H40.11X0, H40.11X1, H40.11X2, H40.11X3, H40.11X4, H40.1210, H40.1211, H40.1212, H40.1213, H40.1214, H40.1220, H40.1221, H40.1222, H40.1223, H40.1224, H40.1230, H40.1231, H40.1232, H40.1233, H40.1234, H40.1290, H40.1291, H40.1292, H40.1293, H40.1294, H40.1310, H40.1311, H40.1312, H40.1313, H40.1314, H40.1320, H40.1321, H40.1322, H40.1323, H40.1324, H40.1330, H40.1331, H40.1332, H40.1333, H40.1334, H40.1390, H40.1391, H40.1392, H40.1393, H40.1394, H40.1410, H40.1411, H40.1412, H40.1413, H40.1414, H40.1420, H40.1421, H40.1422, H40.1423, H40.1424, H40.1430, H40.1431, H40.1432, H40.1433, H40.1434, H40.1490, H40.1491, H40.1492, H40.1493, H40.1494, H40.151, H40.152, H40.153, H40.159, H40.20X0, H40.20X1, H40.20X2, H40.20X3,  H40.20X4, H40.211, H40.212, H40.213, H40.219, H40.2210, H40.2211, H40.2212, H40.2213, H40.2214, H40.2220, H40.2221, H40.2222, H40.2223, H40.2224, H40.2230, H40.2231, H40.2232, H40.2233, H40.2234, H40.2290, H40.2291, H40.2292, H40.2293, H40.2294, H40.231, H40.232, H40.233, H40.239, H40.241, H40.242, H40.243, H40.249, H40.30X0, H40.30X1, H40.30X2, H40.30X3, H40.30X4, H40.31X0, H40.31X1, H40.31X2, H40.31X3, H40.31X4, H40.32X0, H40.32X1, H40.32X2, H40.32X3, H40.32X4, H40.33X0, H40.33X1, H40.33X2, H40.33X3, H40.33X4, H40.40X0, H40.40X1, H40.40X2, H40.40X3, H40.40X4, H40.41X0, H40.41X1, H40.41X2, H40.41X3, H40.41X4, H40.42X0, H40.42X1, H40.42X2, H40.42X3, H40.42X4, H40.43X0, H40.43X1, H40.43X2, H40.43X3, H40.43X4, H40.50X0, H40.50X1, H40.50X2, H40.50X3, H40.50X4, H40.51X0,  H40.51X1, H40.51X2, H40.51X3, H40.51X4, H40.52X0, H40.52X1, H40.52X2, H40.52X3, H40.52X4, H40.53X0, H40.53X1, H40.53X2, H40.53X3, H40.53X4, H40.60X0, H40.60X1, H40.60X2, H40.60X3, H40.60X4, H40.61X0, H40.61X1, H40.61X2, H40.61X3, H40.61X4, H40.62X0, H40.62X1, H40.62X2, H40.62X3, H40.62X4, H40.63X0, H40.63X1, H40.63X2, H40.63X3, H40.63X4, H40.811, H40.812, H40.813, H40.819, H40.821, H40.822, H40.823, H40.829, H40.831, H40.832, H40.833, H40.839, H40.89, H40.9, H42, H44.511, H44.512, H44.513, H44.519, H47.231, H47.232, H47.233, H47.239, Q15.0 (any DX on the claim) |
| Heart Failure and Non-Ischemic Heart Disease | I09.81, I11.0, I13.0, I13.2, I42.0, I42.5, I42.6, I42.7, I42.8, I43, I50.1, I50.20, I50.21, I50.22, I50.23, I50.30, I50.31, I50.32, I50.33, I50.40, I50.41, I50.42, I50.43, I50.810, I50.811, I50.812, I50.813, I50.814, I50.82, I50.83, I50.84, I50.89, I50.9, P29.0 (any DX on the claim) |
| Hip/Pelvic Fracture | M80.051A, M80.052A, M80.059A, M80.0B1A, M80.0B2A, M80.0B9A, M80.851A, M80.852A, M80.859A, M80.8B1A, M80.8B2A, M80.8B9A, M84.350A, M84.351A, M84.352A, M84.353A, M84.359A, M84.451A, M84.452A, M84.453A, M84.459A, M84.550A, M84.551A, M84.552A, M84.553A, M84.559A, M84.650A, M84.651A, M84.652A, M84.653A, M84.659A, M97.01XA, M97.02XA, S32.301A, S32.301B, S32.302A, S32.302B, S32.309A, S32.309B, S32.311A, S32.311B, S32.312A, S32.312B, S32.313A, S32.313B, S32.314A, S32.314B, S32.315A, S32.315B, S32.316A, S32.316B, S32.391A, S32.391B, S32.392A, S32.392B, S32.399A, S32.399B, S32.401A, S32.401B, S32.402A, S32.402B, S32.409A, S32.409B, S32.411A, S32.411B, S32.412A, S32.412B, S32.413A, S32.413B, S32.414A, S32.414B, S32.415A, S32.415B, S32.416A, S32.416B, S32.421A, S32.421B, S32.422A, S32.422B, S32.423A, S32.423B, S32.424A, S32.424B, S32.425A, S32.425B, S32.426A, S32.426B, S32.431A, S32.431B, S32.432A, S32.432B, S32.433A, S32.433B, S32.434A, S32.434B, S32.435A, S32.435B, S32.436A, S32.436B, S32.441A, S32.441B, S32.442A, S32.442B, S32.443A, S32.443B,  S32.444A, S32.444B, S32.445A, S32.445B, S32.446A, S32.446B, S32.451A, S32.451B, S32.452A, S32.452B, S32.453A, S32.453B, S32.454A, S32.454B, S32.455A, S32.455B, S32.456A, S32.456B, S32.461A, S32.461B, S32.462A, S32.462B, S32.463A, S32.463B, S32.464A, S32.464B, S32.465A, S32.465B, S32.466A, S32.466B, S32.471A, S32.471B, S32.472A, S32.472B, S32.473A, S32.473B, S32.474A, S32.474B, S32.475A, S32.475B, S32.476A, S32.476B, S32.481A, S32.481B, S32.482A, S32.482B, S32.483A, S32.483B, S32.484A, S32.484B, S32.485A, S32.485B, S32.486A, S32.486B, S32.491A, S32.491B, S32.492A, S32.492B, S32.499A, S32.499B, S32.501A, S32.501B, S32.502A, S32.502B, S32.509A, S32.509B, S32.511A, S32.511B, S32.512A, S32.512B, S32.519A, S32.519B, S32.591A, S32.591B, S32.592A, S32.592B, S32.599A, S32.599B, S32.601A, S32.601B, S32.602A, S32.602B, S32.609A, S32.609B,  S32.611A, S32.611B, S32.612A, S32.612B, S32.613A, S32.613B, S32.614A, S32.614B, S32.615A, S32.615B, S32.616A, S32.616B, S32.691A, S32.691B, S32.692A, S32.692B, S32.699A, S32.699B, S32.810A, S32.810B, S32.811A, S32.811B, S32.82XA, S32.82XB, S32.89XA, S32.89XB, S32.9XXA, S32.9XXB, S72.001A, S72.001B, S72.001C, S72.002A, S72.002B, S72.002C, S72.009A, S72.009B, S72.009C, S72.011A, S72.011B, S72.011C, S72.012A, S72.012B, S72.012C, S72.019A, S72.019B, S72.019C, S72.021A, S72.021B, S72.021C, S72.022A, S72.022B, S72.022C, S72.023A, S72.023B, S72.023C, S72.024A, S72.024B, S72.024C, S72.025A, S72.025B, S72.025C, S72.026A, S72.026B, S72.026C, S72.031A, S72.031B, S72.031C, S72.032A, S72.032B, S72.032C, S72.033A, S72.033B, S72.033C, S72.034A, S72.034B, S72.034C, S72.035A, S72.035B, S72.035C, S72.036A, S72.036B, S72.036C, S72.041A, S72.041B,  S72.041C, S72.042A, S72.042B, S72.042C, S72.043A, S72.043B, S72.043C, S72.044A, S72.044B, S72.044C, S72.045A, S72.045B, S72.045C, S72.046A, S72.046B, S72.046C, S72.051A, S72.051B, S72.051C, S72.052A, S72.052B, S72.052C, S72.059A, S72.059B, S72.059C, S72.061A, S72.061B, S72.061C, S72.062A, S72.062B, S72.062C, S72.063A, S72.063B, S72.063C, S72.064A, S72.064B, S72.064C, S72.065A, S72.065B, S72.065C, S72.066A, S72.066B, S72.066C, S72.091A, S72.091B, S72.091C, S72.092A, S72.092B, S72.092C, S72.099A, S72.099B, S72.099C, S72.101A, S72.101B, S72.101C, S72.102A, S72.102B, S72.102C, S72.109A, S72.109B, S72.109C, S72.111A, S72.111B, S72.111C, S72.112A, S72.112B, S72.112C, S72.113A, S72.113B, S72.113C, S72.114A, S72.114B, S72.114C, S72.115A, S72.115B, S72.115C, S72.116A, S72.116B, S72.116C, S72.121A, S72.121B, S72.121C, S72.122A, S72.122B,  S72.122C, S72.123A, S72.123B, S72.123C, S72.124A, S72.124B, S72.124C, S72.125A, S72.125B, S72.125C, S72.126A, S72.126B, S72.126C, S72.131A, S72.131B, S72.131C, S72.132A, S72.132B, S72.132C, S72.133A, S72.133B, S72.133C, S72.134A, S72.134B, S72.134C, S72.135A, S72.135B, S72.135C, S72.136A, S72.136B, S72.136C, S72.141A, S72.141B, S72.141C, S72.142A, S72.142B, S72.142C, S72.143A, S72.143B, S72.143C, S72.144A, S72.144B, S72.144C, S72.145A, S72.145B, S72.145C, S72.146A, S72.146B, S72.146C, S72.21XA, S72.21XB, S72.21XC, S72.22XA, S72.22XB, S72.22XC, S72.23XA, S72.23XB, S72.23XC, S72.24XA, S72.24XB, S72.24XC, S72.25XA, S72.25XB, S72.25XC, S72.26XA, S72.26XB, S72.26XC, S79.001A, S79.002A, S79.009A, S79.011A, S79.012A, S79.019A, S79.091A, S79.092A, S79.099A (any DX on the claim) |
| Hyperlipidemia | E78.0, E78.00, E78.01, E78.1, E78.2, E78.3, E78.4, E78.41, E78.49, E78.5 (any DX on the claim) |
| Hypertension | H35.031, H35.032, H35.033, H35.039, I10, I11.0, I11.9, I12.0, I12.9, I13.0, I13.10, I13.11, I13.2, I15.0, I15.1, I15.2, I15.8, I15.9, I1A.0, I67.4, N26.2 (any DX on the claim) |
| Hypothyroidism | E00.0, E00.1, E00.2, E00.9, E01.8, E02, E03.0, E03.1, E03.2, E03.3, E03.4, E03.8, E03.9, E89.0 (any DX on the claim) |
| Ischemic Heart Disease | I20.0, I20.1, I20.2, I20.8, I20.81, I20.89, I24.0, I24.1, I24.8, I24.81, I24.89, I25.10, I25.110, I25.111, I25.112, I25.118, I25.119, I25.3, I25.41, I25.42, I25.5, I25.6, I25.700, I25.701, I25.702, I25.708, I25.710, I25.711, I25.712, I25.718, I25.719, I25.720, I25.721, I25.722, I25.728, I25.729, I25.730, I25.731, I25.732, I25.738, I25.739, I25.750, I25.751, I25.752, I25.758, I25.759, I25.760, I25.761, I25.762, I25.768, I25.769, I25.790, I25.791, I25.792, I25.798, I25.799, I25.810, I25.811, I25.812, I25.82, I25.83, I25.84, I25.85, I25.89, I25.9 (any DX on the claim) |
| Non-Alzheimer's Dementia† | F01.50, F01.51, F01.511, F01.518, F01.52, F01.53, F01.54, F01.A0, F01.A11, F01.A18, F01.A2, F01.A3, F01.A4, F01.B0, F01.B11, F01.B18, F01.B2, F01.B3, F01.B4, F01.C0, F01.C11, F01.C18, F01.C2, F01.C3, F01.C4, F02.80, F02.81, F02.811, F02.818, F02.82, F02.83, F02.84, F02.A0, F02.A11, F02.A18, F02.A2, F02.A3, F02.A4, F02.B0, F02.B11, F02.B18, F02.B2, F02.B3, F02.B4, F02.C0,  F02.C11, F02.C18, F02.C2, F02.C3, F02.C4, F03.90, F03.91, F03.911, F03.918, F03.92, F03.93, F03.94, F03.A0, F03.A11, F03.A18, F03.A2, F03.A3, F03.A4, F03.B0, F03.B11, F03.B18, F03.B2, F03.B3, F03.B4, F03.C0, F03.C11, F03.C18, F03.C2, F03.C3, F03.C4, F05, G13.8, G31.01, G31.09, G31.1, G31.2, G31.83, G94, R41.81 (any DX on the claim) |
| Osteoporosis With or Without Pathological Fracture | M80.00XA, M80.011A, M80.012A, M80.019A, M80.021A, M80.022A, M80.029A, M80.031A, M80.032A, M80.039A, M80.041A, M80.042A, M80.049A, M80.051A, M80.052A, M80.059A, M80.061A, M80.062A, M80.069A, M80.071A, M80.072A, M80.079A, M80.08XA, M80.0AXA, M80.0B1A, M80.0B2A, M80.0B9A, M80.80XA, M80.811A, M80.812A, M80.819A, M80.821A, M80.822A, M80.829A, M80.831A, M80.832A, M80.839A, M80.841A, M80.842A, M80.849A, M80.851A, M80.852A, M80.859A, M80.861A, M80.862A, M80.869A, M80.871A, M80.872A, M80.879A, M80.88XA, M80.8AXA, M80.8B1A, M80.8B2A, M80.8B9A, M81.0,  M81.6, M81.8 (any DX on the claim) |
| Parkinson's Disease and Secondary Parkinsonism | G20, G20.A1, G20.A2, G20.B1, G20.B2, G20.C, G21.11, G21.19, G21.3, G21.4, G21.8, G21.9, G31.83 (any DX on the claim) |
| Pneumonia, All-cause | A01.03, A02.22, A06.5, A20.2, A21.2, A22.1, A31.0, A37.01, A37.11, A37.81, A37.91, A40.3, A42.0, A43.0, A48.1, A50.04, A54.84, B01.2, B05.2, B06.81, B37.1, B38.0, B38.2, B39.0, B39.2, B40.0, B40.2, B41.0, B58.3, B59, B66.4, B67.1, B77.81, B95.3, B96.0, B96.1, J09.X1, J10.00, J10.01, J10.08, J11.00, J11.08, J12.0, J12.1, J12.2, J12.3, J12.81, J12.82, J12.89, J12.9, J13, J14, J15.0, J15.1, J15.20, J15.211, J15.212, J15.29, J15.3, J15.4, J15.5, J15.6, J15.61, J15.69, J15.7, J15.8, J15.9, J16.0, J16.8, J17, J18.0, J18.1, J18.2, J18.8, J18.9, J20.0, J84.111, J84.116, J84.117, J84.178, J84.2, J85.1, J95.851, P23.0, P23.1, P23.2, P23.3, P23.4, P23.5, P23.6, P23.8, P23.9, Z87.01 (any DX on the claim) |
| Rheumatoid Arthritis/Osteoarthritis | L40.50, L40.51, L40.54, L40.59, M05.00, M05.011, M05.012, M05.019, M05.021, M05.022, M05.029, M05.031, M05.032, M05.039, M05.041, M05.042, M05.049, M05.051, M05.052, M05.059, M05.061, M05.062, M05.069, M05.071, M05.072,  M05.079, M05.09, M05.10, M05.111, M05.112, M05.119, M05.121, M05.122, M05.129, M05.131, M05.132, M05.139, M05.141, M05.142, M05.149, M05.151, M05.152, M05.159, M05.161, M05.162, M05.169, M05.171, M05.172, M05.179,  M05.19, M05.20, M05.211, M05.212, M05.219, M05.221, M05.222, M05.229, M05.231, M05.232, M05.239, M05.241, M05.242, M05.249, M05.251, M05.252, M05.259, M05.261, M05.262, M05.269, M05.271, M05.272, M05.279, M05.29,  M05.30, M05.311, M05.312, M05.319, M05.321, M05.322, M05.329, M05.331, M05.332, M05.339, M05.341, M05.342, M05.349, M05.351, M05.352, M05.359, M05.361, M05.362, M05.369, M05.371, M05.372, M05.379, M05.39, M05.40,  M05.411, M05.412, M05.419, M05.421, M05.422, M05.429, M05.431, M05.432, M05.439, M05.441, M05.442, M05.449, M05.451, M05.452, M05.459, M05.461, M05.462, M05.469, M05.471, M05.472, M05.479, M05.49, M05.50, M05.511,  M05.512, M05.519, M05.521, M05.522, M05.529, M05.531, M05.532, M05.539, M05.541, M05.542, M05.549, M05.551, M05.552, M05.559, M05.561, M05.562, M05.569, M05.571, M05.572, M05.579, M05.59, M05.60, M05.611, M05.612,  M05.619, M05.621, M05.622, M05.629, M05.631, M05.632, M05.639, M05.641, M05.642, M05.649, M05.651, M05.652, M05.659, M05.661, M05.662, M05.669, M05.671, M05.672, M05.679, M05.69, M05.70, M05.711, M05.712, M05.719,  M05.721, M05.722, M05.729, M05.731, M05.732, M05.739, M05.741, M05.742, M05.749, M05.751, M05.752, M05.759, M05.761, M05.762, M05.769, M05.771, M05.772, M05.779, M05.79, M05.7A, M05.80, M05.811, M05.812, M05.819,  M05.821, M05.822, M05.829, M05.831, M05.832, M05.839, M05.841, M05.842, M05.849, M05.851, M05.852, M05.859, M05.861, M05.862, M05.869, M05.871, M05.872, M05.879, M05.89, M05.8A, M05.9, M06.00, M06.011, M06.012, M06.019,  M06.021, M06.022, M06.029, M06.031, M06.032, M06.039, M06.041, M06.042, M06.049, M06.051, M06.052, M06.059, M06.061, M06.062, M06.069, M06.071, M06.072, M06.079, M06.08, M06.09, M06.0A, M06.1, M06.20, M06.211, M06.212,  M06.219, M06.221, M06.222, M06.229, M06.231, M06.232, M06.239, M06.241, M06.242, M06.249, M06.251, M06.252, M06.259, M06.261, M06.262, M06.269, M06.271, M06.272, M06.279, M06.28, M06.29, M06.30, M06.311, M06.312, M06.319, M06.321, M06.322, M06.329, M06.331, M06.332, M06.339, M06.341, M06.342, M06.349, M06.351, M06.352, M06.359, M06.361, M06.362, M06.369, M06.371, M06.372, M06.379, M06.38, M06.39, M06.80, M06.811, M06.812, M06.819, M06.821, M06.822, M06.829, M06.831, M06.832, M06.839, M06.841, M06.842, M06.849, M06.851, M06.852, M06.859, M06.861, M06.862, M06.869, M06.871, M06.872, M06.879, M06.88, M06.89, M06.8A, M06.9, M08.00, M08.011, M08.012, M08.019, M08.021, M08.022, M08.029, M08.031, M08.032, M08.039, M08.041, M08.042, M08.049, M08.051, M08.052, M08.059, M08.061, M08.062, M08.069, M08.071, M08.072, M08.079, M08.08, M08.09, M08.0A, M08.1, M08.20, M08.211, M08.212, M08.219, M08.221, M08.222, M08.229, M08.231, M08.232, M08.239, M08.241, M08.242, M08.249, M08.251, M08.252, M08.259, M08.261, M08.262, M08.269, M08.271, M08.272, M08.279, M08.28, M08.29, M08.2A, M08.3, M08.40, M08.411, M08.412, M08.419, M08.421, M08.422, M08.429, M08.431, M08.432, M08.439, M08.441, M08.442, M08.449, M08.451, M08.452, M08.459, M08.461, M08.462, M08.469, M08.471, M08.472, M08.479, M08.48, M08.4A, M08.80, M08.811, M08.812, M08.819, M08.821, M08.822, M08.829, M08.831, M08.832, M08.839, M08.841, M08.842, M08.849, M08.851, M08.852, M08.859, M08.861, M08.862, M08.869, M08.871, M08.872, M08.879, M08.88, M08.89, M08.90, M08.911, M08.912, M08.919, M08.921, M08.922, M08.929, M08.931, M08.932, M08.939, M08.941, M08.942, M08.949, M08.951, M08.952, M08.959, M08.961, M08.962, M08.969, M08.971, M08.972, M08.979, M08.98, M08.99, M08.9A, M15.0, M15.1, M15.2, M15.3, M15.4, M15.8, M15.9, M16.0, M16.10, M16.11, M16.12, M16.2, M16.30, M16.31, M16.32, M16.4, M16.50, M16.51,  M16.52, M16.6, M16.7, M16.9, M17.0, M17.10, M17.11, M17.12, M17.2, M17.30, M17.31, M17.32, M17.4, M17.5, M17.9, M18.0, M18.10, M18.11, M18.12, M18.2, M18.30, M18.31, M18.32, M18.4, M18.50, M18.51, M18.52, M18.9, M19.011,  M19.012, M19.019, M19.021, M19.022, M19.029, M19.031, M19.032, M19.039, M19.041, M19.042, M19.049, M19.071, M19.072, M19.079, M19.09, M19.111, M19.112, M19.119, M19.121, M19.122, M19.129, M19.131, M19.132, M19.139,  M19.141, M19.142, M19.149, M19.171, M19.172, M19.179, M19.19, M19.211, M19.212, M19.219, M19.221, M19.222, M19.229, M19.231, M19.232, M19.239, M19.241, M19.242, M19.249, M19.271, M19.272, M19.279, M19.29, M19.90, M19.91, M19.92, M19.93, M45.0, M45.1, M45.2, M45.3, M45.4, M45.5, M45.6, M45.7, M45.8, M45.9, M45.A0, M45.A1, M45.A2, M45.A3, M45.A4, M45.A5, M45.A6, M45.A7, M45.A8, M45.AB, M46.80, M46.81, M46.82, M46.83, M46.84, M46.85, M46.86, M46.87, M46.88, M46.89, M46.90, M46.91, M46.92, M46.93, M46.94, M46.95, M46.96, M46.97, M46.98, M46.99, M47.011, M47.012, M47.013, M47.014, M47.015, M47.016, M47.019, M47.021, M47.022, M47.029, M47.10, M47.11, M47.12, M47.13, M47.14, M47.15, M47.16, M47.20, M47.21, M47.22, M47.23, M47.24, M47.25, M47.26, M47.27, M47.28, M47.811, M47.812, M47.813, M47.814, M47.815, M47.816, M47.817, M47.818, M47.819, M47.891, M47.892, M47.893, M47.894, M47.895, M47.896, M47.897, M47.898, M47.899, M47.9, M48.8X1, M48.8X2, M48.8X3, M48.8X4, M48.8X5, M48.8X6, M48.8X7,  M48.8X8, M48.8X9 (any DX on the claim) |
| Stroke/Transient Ischemic Attack | G45.0, G45.1, G45.2, G45.3, G45.8, G45.9, G46.0, G46.1, G46.2, G46.3, G46.4, G46.5, G46.6, G46.7, G46.8, G97.31, G97.32, I60.00, I60.01, I60.02, I60.10, I60.11, I60.12, I60.2, I60.20, I60.21, I60.22, I60.30, I60.31, I60.32, I60.4, I60.50, I60.51, I60.52, I60.6, I60.7, I60.8, I60.9, I61.0, I61.1, I61.2, I61.3, I61.4, I61.5, I61.6, I61.8, I61.9, I62.00, I62.01, I62.02, I62.9, I63.00, I63.011, I63.012, I63.013, I63.019, I63.02, I63.031, I63.032, I63.033, I63.039, I63.09, I63.10, I63.111, I63.112, I63.113, I63.119, I63.12, I63.131, I63.132, I63.133, I63.139, I63.19, I63.20, I63.211, I63.212, I63.213, I63.219, I63.22, I63.231, I63.232, I63.233, I63.239, I63.29, I63.30, I63.311, I63.312, I63.313, I63.319, I63.321, I63.322, I63.323, I63.329, I63.331, I63.332, I63.333, I63.339, I63.341, I63.342, I63.343, I63.349, I63.39, I63.40, I63.411, I63.412, I63.413, I63.419, I63.421, I63.422, I63.423, I63.429, I63.431, I63.432, I63.433, I63.439, I63.441, I63.442, I63.443, I63.449, I63.49, I63.50, I63.511, I63.512, I63.513, I63.519, I63.521, I63.522, I63.523, I63.529, I63.531, I63.532, I63.533, I63.539, I63.541, I63.542, I63.543, I63.549, I63.59, I63.6, I63.8, I63.81, I63.89, I63.9, I67.841, I67.848, I67.89, I97.810, I97.811, I97.820, I97.821 (any DX on the claim)  EXCLUSION: If any of the qualifying claims have any of the following codes in any DX position then EXCLUDE: S06.340A, S06.341A, S06.342A, S06.343A, S06.344A, S06.345A, S06.346A, S06.347A, S06.348A, S06.34AA, S06.349A, S06.350A, S06.351A, S06.352A, S06.353A, S06.354A, S06.355A, S06.356A, S06.357A, S06.358A, S06.35AA, S06.359A, S06.360A, S06.361A, S06.362A, S06.363A, S06.364A, S06.365A, S06.366A, S06.367A, S06.368A, S06.36AA, S06.369A, S06.370A, S06.371A, S06.372A, S06.373A, S06.374A, S06.375A, S06.376A, S06.377A, S06.378A, S06.37AA, S06.379A, S06.380A, S06.381A, S06.382A, S06.383A, S06.384A, S06.385A, S06.386A, S06.387A, S06.388A, S06.38AA, S06.389A, S06.5X0A, S06.5X1A, S06.5X2A, S06.5X3A, S06.5X4A, S06.5X5A,  S06.5X6A, S06.5X7A, S06.5X8A, S06.5XAA, S06.5X9A, S06.6X0A, S06.6X1A, S06.6X2A, S06.6X3A, S06.6X4A, S06.6X5A, S06.6X6A, S06.6X7A, S06.6X8A, S06.6XAA, S06.6X9A, S06.810A, S06.811A, S06.812A, S06.813A, S06.814A, S06.815A, S06.816A, S06.817A, S06.818A, S06.81AA, S06.819A, S06.820A, S06.821A, S06.822A, S06.823A, S06.824A, S06.825A, S06.826A, S06.827A, S06.828A, S06.82AA, S06.829A, S06.890A, S06.891A, S06.892A, S06.893A, S06.894A, S06.895A, S06.896A, S06.897A, S06.898A, S06.89AA, S06.899A, S06.9X0A, S06.9X1A, S06.9X2A, S06.9X3A, S06.9X4A, S06.9X5A, S06.9X6A, S06.9X7A, S06.9X8A, S06.9XAA, S06.9X9A, S06.A0XA, S06.A1XA |

| **Medication variables** | **Generic drug names** |
| --- | --- |
| Antiplatelet agents | Clopidogrel, Ticagrelor, Ticlopidine, Prasugrel |
| Oral anticoagulants | Apixaban, Betrixaban, Dabigatran etexilate, Edoxaban, Rivaroxaban |
| Antihypertensives | ACEIs, ARBs, Beta blockers, CCB, diuretic, Aldosterone receptor antagonists, Antihypertensives |
| statins | Lovastatin, Rosuvastatin, Fluvastatin, atorvastatin, Pitavastatin, Lovastatin, Pravastatin, Simvastatin |
| NSAIDs | Amlodipine-celecoxib, Celecoxib, Diclofenac, Diclofenac topical, Diclofenac-misoprostol, Esomeprazole-naproxen, Etodolac, Famotidine-ibuprofen, Fenoprofen, Flurbiprofen, Ibuprofen, Indomethacin, Ketoprofen, Ketorolac, Meclofenamate, Mefenamic acid, Meloxicam, Nabumetone, Naproxen, Naproxen-sumatriptan, Oxaprozin, Piroxicam, Sulindac, Tolmetin |
| Opioids | Codeine, Fentanyl, Hydromorphone, Meperidine, Methadone, Morphine, Oxycodone, Tramadol, Hydrocodone, Buprenorphine |
| Antidepressants | Amitriptyline Hcl, Amitriptyline/Chlordiazepoxide, Amoxapine, Bupropion Hcl, Bupropion Hbr, Citalopram Hydrobromide, Clomipramine Hcl, Desipramine Hcl, Doxepin Hcl, Duloxetine Hcl, Desvenlafaxine, Desvenlafaxin Succinate, Escitalopram Oxalate, Fluoxetine Hcl, Fluvoxamine Maleate, Imipramine Hcl, Imipramine Pamoate, Isocarboxazid, Levomilnacipran Hcl, Lurasidone Hcl, Mirtazapine, Nefazodone Hcl, Nortriptyline Hcl, Olanzapine, Olanzapine/Fluoxetine Hcl, Phenelzine Sulfate, Perphenazine/Amitriptyline Hcl, Protriptyline Hcl, Paroxetine Hcl, Paroxetine Mesylate, Quetiapine Fumarate, Sertraline Hcl, Selegiline, Tranylcypromine Sulfate, Trimipramine Maleate, Trazodone Hcl, Vortioxetine Hydrobromide, Vilazodone Hcl, Venlafaxine Hcl |
| Antipsychotics | Chlorpromazine, Droperidol, Fluphenazine, Haloperidol, Loxapine, Lithium, Mesoridazine, Methotrimeprazine, Molindone, Perphenazine, Pimozide, Prochlorperazine, Promazine, Thioridazine, Thiothixene, Trifluoperazine, Triflupromazine, Aripiprazole, Asenapine, Cariprazine, Clozapine, Iloperidone, Lumateperone, Lurasidone, Olanzapine, Paliperidone, Quetiapine, Risperidone, Ziprasidone |
| Anti-dementia | Donepezil, Memantine, Rivastigmine, Galantamine, Aducanumab |
| PPIs | Dexlansoprazole, Esomeprazole, Lansoprazole, Omeprazole, Omeprazole-sodium bicarbonate, Pantoprazole, Rabeprazole |
| GLDs | SGLT, Sulfonylurea, GLP, Thiazolidinedione, DPP4i, Alpha glucosidase inhibitor,Meglitinide |
| Antidiabetics | Insulin Degludec, Insulin Glulisine, Human, Insulin Glargine, Insulin Lispro, Insulin Detemir, Insulin, Regular, Human, Insulin Aspart, Human, Acarbose, Albiglutide, Alogliptin, Canagliflozin, Chlorpropamide, Dapagliflozin, Exenatide, Glimepiride, Glipizide, Glyburide (Glibenclamide), Linagliptin, Liraglutide, Metformin, Miglitol, Nateglinide,Pioglitazone, Repaglinide, Rosiglitazone, Saxagliptin, Sitagliptin, Tolazamide, Tolbutamide, Troglitazone |

**Table S3. Definitions of the candidate predictors for EHR care-continuity**

| **Predictors** | **Definitions** |
| --- | --- |
| Having general medical exam or routine care* | ICD-9: V70.0 or V72.3 |
| Mammography* | ICD9: V76.12  CPT: 76082 76083 76092  HCPS: G0202 G0203 |
| Pap smear* | HCPCS: Q0091 P3000 P3001 G0123 G0124 G0141 G0143 G0145 G0147  G0148  CPT: 88141 – 88158 88164 – 88167  88174 – 88175 |
| PSA Test* | HCPCS: G0103  CPT: 84153, 84152 – 84154 |
| Colonoscopy* | ICD-9: 45.23  HCPCS: G0105 G0121  CPT: 45378-45392 |
| Fecal occult blood test* | HCPCS:G0107 G0328  CPT:82270 82274 |
| Influenza vaccine* | CPT: 90655 – 90660 90724  HCPCS: G0008  ICD-9: V04.8 V04.81 V06.6 |
| Pneumococcal vaccine* | CPT: 90669 90670 90732  ICD-9: V03.82 |
| Having BMI recorded* | With BMI recorded in the EHR |
| Having A1C ordered or value recorded* | Ordering or with results available in the EHR for HbA1c |
| Having 2 of the above routine care facts** | As described |
| With any one medication use record | As described |
| With at least 2 medication use records | As described |
| With 1 diagnosis recorded in the EHR | As described |
| With at least 2 diagnoses recorded in the EHR | As described |
| Having at least one inpatient or outpatient encounter in EHR | As described |
| Having at least two outpatient encounters in the EHR | As described |
| Having ED visit in the EHR | As described |
| Having seen the same provider twice | As described |
| Having seen the same provider >=3 times | As described |

^**^having 2 of the facts followed by *, EHR = electronic health records, PSA = prostate specific antigen

**Table S4. Population characteristics of training, testing, and validation cohort**

|  | **OneFlorida+** | | **REACHnet–LABlue** |
| --- | --- | --- | --- |
| **Variable** | **Training Cohort (N)** | **Testing Cohort (N)** | **Validation Cohort (N)** |
| **Total** | 5462 (100.0)% | 3364 (100.0)% | 14567 (100%) |
| **AGE_AT_INDEX** | 47.03 (15.61) | 48.49 (14.2) | 45.14 (13.16) |
| **BMI** | | | |
| Mean (SD) | 31.48 (8.94) | 32.16 (9.04) | 29.55 (7.11) |
| BMI_missing | 973 (17.81)% | 587 (17.45)% | 1389 (9.54)% |
| **Blood pressure** | | | |
| Systolic blood pressure Mean (SD) | 128.42 (19.36) | 130.12 (19.6) | 124.67 (13.82) |
| Systolic blood pressure_missing | 1075 (19.68)% | 655 (19.47)% | 1535 (10.54)% |
| Diastolic blood pressure Mean (SD) | 76.3 (11.86) | 77.41 (11.56) | 77.7(8.66) |
| Diastolic blood pressure_missing | 1074 (19.66)% | 655 (19.47)% | 1535 (10.54)% |
| **Race_Ethnicity** | | | |
| Hispanics | 1437 (26.31)% | 855 (25.42)% | 491 (3.37)% |
| NHB | 1706 (31.23)% | 1005 (29.88)% | 2256 (15.49)% |
| NHW | 2159 (39.53)% | 1382 (41.08)% | 11408 (78.31)% |
| Others | 43 (0.79)% | 33 (0.98)% | 251 (1.72)% |
| Unknown | 117 (2.14)% | 89 (2.65)% | 161 (1.11)% |
| **SEX** | | | |
| SEX_F | 3649 (66.81)% | 2232 (66.35)% | 9437 (64.78)% |
| **Smoking_Status** | | | |
| Current Smoker | 946 (17.32)% | 519 (15.43)% | 626 (4.3)% |
| Former Smoker | 653 (11.96)% | 521 (15.49)% | 736 (5.05)% |
| Never Smoker | 1246 (22.81)% | 790 (23.48)% | 11808 (81.06)% |
| Unknown | 2617 (47.91)% | 1534 (45.6)% | 1397 (9.59)% |
| **Labs** | | | |
| HbA1c Mean (SD) | 7.01 (1.85) | 7.18 (2.04) | 5.53 (0.93) |
| HbA1c_missing | 4053 (74.2)% | 2350 (69.86)% | 9074 (62.29)% |
| HDL Mean (SD) | 46.53 (16.08) | 46.59 (15.0) | 55.28 (15.43) |
| HDL_missing | 4187 (76.66)% | 2484 (73.84)% | 6498 (44.61)% |
| LDL Mean (SD) | 96.08 (43.6) | 82.67 (27.59) | 108.78 (32.54) |
| LDL_missing | 5378 (98.46)% | 3358 (99.82)% | 14251 (97.83)% |
| Triglycerides Mean (SD) | 163.01 (152.57) | 160.07 (175.2) | 111.65 (67.15) |
| Triglycerides_missing | 4182 (76.57)% | 2480 (73.72)% | 6488 (44.54)% |
| GGT Mean (SD) | 121.07 (164.57) | 110.86 (131.86) | 189.7 (35.38) |
| GGT_missing | 5406 (98.97)% | 3321 (98.72)% | 6496 (44.59)% |
| **Common and CCI disease** | | | |
| Obesity | 1273 (23.31)% | 950 (28.24)% | 2161 (14.83)% |
| Hypertension | 2361 (43.23)% | 1543 (45.87)% | 1957 (13.43)% |
| Stroke/Transient Ischemic Attack | 183 (3.35)% | 111 (3.3)% | 70 (0.48)% |
| Myocardial infarction | 215 (3.94)% | 125 (3.72)% | 59 (0.41)% |
| Ischemic Heart Disease | 488 (8.93)% | 307 (9.13)% | 244 (1.68)% |
| Family history of diabetes | 288 (5.27)% | 126 (3.75)% | 60 (0.41)% |
| Alcohol use disorder | 219 (4.01)% | 120 (3.57)% | 65 (0.45)% |
| Type 2 Diabetes (T2D) | 1806 (33.06)% | 1277 (37.96)% | 715 (4.91)% |
| Acute Myocardial Infarction | 69 (1.26)% | 46 (1.37)% | 31 (0.21)% |
| Congestive heart failure | 421 (7.71)% | 296 (8.8)% | 95 (0.65)% |
| Peripheral vascular disorder | 407 (7.45)% | 280 (8.32)% | 198 (1.36)% |
| Cerebrovascular disease | 338 (6.19)% | 203 (6.03)% | 133 (0.91)% |
| Dementia | 48 (0.88)% | 28 (0.83)% | 8 (0.05)% |
| Chronic pulmonary disease | 1040 (19.04)% | 605 (17.98)% | 751 (5.16)% |
| Rheumatoid arthritis/collagen vascular diseases | 239 (4.38)% | 188 (5.59)% | 946 (6.49)% |
| Ulcer disease | 63 (1.15)% | 37 (1.1)% | 40 (0.27)% |
| Mild liver disease | 434 (7.95)% | 285 (8.47)% | 394 (2.7)% |
| Uncomplicated diabetes | 1555 (28.47)% | 1008 (29.96)% | 550 (3.78)% |
| Complicated diabetes | 957 (17.52)% | 851 (25.3)% | 429 (2.95)% |
| Hemiplegia or paraplegia | 127 (2.33)% | 58 (1.72)% | 29 (0.2)% |
| Renal failure | 425 (7.78)% | 293 (8.71)% | 177 (1.22)% |
| Any tumor | 573 (10.49)% | 376 (11.18)% | 415 (2.85)% |
| Moderate or severe liver disease | 66 (1.21)% | 40 (1.19)% | 17 (0.12)% |
| Metastatic cancer | 171 (3.13)% | 115 (3.42)% | 64 (0.44)% |
| AIDS/HIV | 142 (2.6)% | 60 (1.78)% | 63 (0.43)% |
| **Providers** | | | |
| Behavioral Health & Social Service Providers | 71 (1.3)% | 90 (2.68)% | 430 (2.95)% |
| Chiropractic Providers | 0 (0.0)% | 0 (0.0)% | 224 (1.54)% |
| Dental Providers | 37 (0.68)% | 9 (0.27)% | 31 (0.21)% |
| Dietary & Nutritional Service Providers | 10 (0.18)% | 33 (0.98)% | 273 (1.87)% |
| Emergency Medical Service Providers | 0 (0.0)% | 0 (0.0)% | 0 (0)% |
| Eye and Vision Services Providers | 85 (1.56)% | 80 (2.38)% | 1062 (7.29)% |
| Nursing Service Providers | 214 (3.92)% | 338 (10.05)% | 255 (1.75)% |
| Other Service Providers | 48 (0.88)% | 40 (1.19)% | 517 (3.55)% |
| Pharmacy Service Providers | 19 (0.35)% | 3 (0.09)% | 8 (0.05)% |
| Group | 8 (0.15)% | 2 (0.06)% | 0 (0)% |
| Allopathic & Osteopathic Physicians | 5070 (92.82)% | 3026 (89.95)% | 14073 (96.61)% |
| Podiatric Medicine & Surgery Service Providers | 132 (2.42)% | 101 (3.0)% | 970 (6.66)% |
| Respiratory, Developmental, Rehabilitative and Restorative Service Providers | 35 (0.64)% | 24 (0.71)% | 97 (0.67)% |
| Speech, Language and Hearing Service Providers | 21 (0.38)% | 7 (0.21)% | 439 (3.01)% |
| Technologists, Technicians & Other Technical Service Providers | 10 (0.18)% | 3 (0.09)% | 127 (0.87)% |
| Agencies | 3 (0.05)% | 3 (0.09)% | 0 (0)% |
| Ambulatory Health Care Facilities | 6 (0.11)% | 4 (0.12)% | 366 (2.51)% |
| Hospital Units | 0 (0.0)% | 0 (0.0)% | 0 (0)% |
| Hospitals | 6 (0.11)% | 0 (0.0)% | 0 (0)% |
| Laboratories | 0 (0.0)% | 0 (0.0)% | 0 (0)% |
| Managed Care Organizations | 0 (0.0)% | 0 (0.0)% | 0 (0)% |
| Nursing & Custodial Care Facilities | 0 (0.0)% | 0 (0.0)% | 0 (0)% |
| Residential Treatment Facilities | 0 (0.0)% | 0 (0.0)% | 0 (0)% |
| Suppliers | 0 (0.0)% | 0 (0.0)% | 0 (0)% |
| Transportation Services | 0 (0.0)% | 0 (0.0)% | 0 (0)% |
| Physician Assistants & Advanced Practice Nursing Providers | 2056 (37.64)% | 1682 (50.0)% | 10148 (69.66)% |
| Nursing Service Related Providers | 14 (0.26)% | 2 (0.06)% | 0 (0)% |
| Respite Care Facility | 0 (0.0)% | 0 (0.0)% | 0 (0)% |
| Student, Health Care | 352 (6.44)% | 320 (9.51)% | 1917 (13.16)% |
| Others | 0 (0.0)% | 0 (0.0)% | 0 (0)% |
| **Outpatient Encounter_Count Mean (SD)** | 8.58 (11.74) | 7.97 (10.78) | 11.4 (13.98) |
| **Inpatient Encounter_Count Mean (SD)** | 0.28 (0.97) | 0.19 (0.75) | 0.04 (0.23) |
| **Same Provider Twice** | 667 (12.21)% | 438 (13.02)% | 5191 (35.64)% |
| **Same Provider >=3** | 4618 (84.55)% | 2973 (88.38)% | 9039 (62.05)% |
| **Influenza vaccine** | 247 (4.52)% | 157 (4.67)% | 0 (0)% |
| **Pneumococcal vaccine** | 90 (1.65)% | 32 (0.95)% | 0 (0)% |
| **Mammography** | 206 (3.77)% | 306 (9.1)% | 941 (6.46)% |
| **Pap smear** | 36 (0.66)% | 24 (0.71)% | 3402 (23.35)% |
| **Colonoscopy** | 268 (4.91)% | 191 (5.68)% | 1876 (12.88)% |
| **PSA Test** | 45 (0.82)% | 44 (1.31)% | 127 (0.87)% |
| **Fecal occult blood test** | 89 (1.63)% | 51 (1.52)% | 23 (0.16)% |
| **General medical exam** | 624 (11.42)% | 447 (13.29)% | 154 (1.06)% |
| **Having BMI records** | 4489 (82.19)% | 2777 (82.55)% | 11.4 (13.98) |
| **Having 2 of the above routine care facts** | 792 (14.5)% | 592 (17.6)% | 0.04 (0.23) |

**Table S5. Mean difference and 95% confidence interval by each HEPS decile in CCI score, ECI score, and Combined score in OneFlorida+**

| **HEPS deciles** | **CCI** | **ECI** | **Combined Score** |
| --- | --- | --- | --- |
| 0~<10% | 2.24 [2.1469, 2.3283] | 3.14 [2.8894, 3.3807] | 1.46 [1.3897, 1.5268] |
| 10~<20% | 1.34 [1.2357, 1.4387] | 1.69 [1.4124, 1.9733] | 0.89 [0.8107, 0.9657] |
| 20~<30% | 0.83 [0.7294, 0.9268] | 1.14 [0.8461, 1.4333] | 0.54 [0.4598, 0.6115] |
| 30~<40% | 0.48 [0.3933, 0.5653] | 0.76 [0.4792, 1.0367] | 0.33 [0.2649, 0.4039] |
| 40~<50% | 0.38 [0.2984, 0.4626] | 0.52 [0.2908, 0.7501] | 0.2 [0.1433, 0.2497] |
| 50~<60% | 0.44 [0.2787, 0.6004] | 0.43 [-0.0176, 0.8748] | 0.26 [0.1546, 0.3619] |
| 60~<70% | 0.41 [0.2081, 0.6162] | 0.36 [-0.238, 0.9677] | 0.24 [0.0857, 0.3873] |
| 70~<80% | 0.39 [0.2053, 0.5747] | -0.13 [-0.7386, 0.4786] | 0.21 [0.1034, 0.3166] |
| >=80% | 0.34 [0.2114, 0.4606] | 0.11 [-0.2029, 0.4296] | 0.15 [0.0823, 0.2254] |

CCI = Charlson Comorbidity Index; ECI = Elixhauser Comorbidity Index

**Table S6. Mean standardized difference and 95% confidence interval by each HEPS decile in 42 selected variables in OneFlorida+**

| **HEPS deciles** | **30 Comorbidity Variables** | **12 Medication Variables** |
| --- | --- | --- |
| 0~<10% | 0.29 [0.2118, 0.3755] | 0.32 [0.2358, 0.4108] |
| 10~<20% | 0.17 [0.1158, 0.2182] | 0.22 [0.1589, 0.2811] |
| 20~<30% | 0.11 [0.0788, 0.1472] | 0.15 [0.1058, 0.2008] |
| 30~<40% | 0.07 [0.0468, 0.0879] | 0.11 [0.0766, 0.1417] |
| 40~<50% | 0.06 [0.0432, 0.0775] | 0.07 [0.0501, 0.0949] |
| 50~<60% | 0.06 [0.0397, 0.0797] | 0.09 [0.0577, 0.124] |
| 60~<70% | 0.05 [0.0241, 0.0679] | 0.08 [0.0589, 0.1061] |
| 70~<80% | 0.05 [0.0263, 0.0683] | 0.09 [0.0466, 0.13] |
| >=80% | 0.04 [0.0257, 0.0583] | 0.08 [0.0506, 0.1027] |

**Table S7. Fairness assessment across race-ethnicity groups in XGBoost models**

| **Complete predictor set** | | | | | | | | | | | | | | | |
| --- | --- | --- | --- | --- | --- | --- | --- | --- | --- | --- | --- | --- | --- | --- | --- |
|  |  | PPV | | FPR | | TPR | | NPV | | te | | FNR | | ACC | |
|  |  | protected | privileged | protected | privileged | protected | privileged | protected | privileged | protected | privileged | protected | privileged | protected | privileged |
| NHB & NHW | mean | 0.48 | 0.42 | 0.36 | 0.30 | 0.75 | 0.65 | 0.85 | 0.86 | 0.31 | 0.40 | 0.25 | 0.35 | 0.68 | 0.69 |
|  | Ratio | 1.14 | | 1.20 | | 1.15 | | 1.00 | | 0.78 | | 0.71 | | 0.98 | |
| H & NHW | mean | 0.49 | 0.43 | 0.11 | 0.29 | 0.40 | 0.65 | 0.85 | 0.86 | 1.46 | 0.41 | 0.60 | 0.35 | 0.79 | 0.69 |
|  | Ratio | 1.16 | | 0.37 | | 0.62 | | 0.99 | | 3.59 | | 1.69 | | 1.14 | |
| **Predictor set identified from Huang et al.** | | | | | | | | | | | | | | | |
|  |  | PPV | | FPR | | TPR | | NPV | | te | | FNR | | ACC | |
|  |  | protected | privileged | protected | privileged | protected | privileged | protected | privileged | protected | privileged | protected | privileged | protected | privileged |
| NHB & NHW | mean | 0.48 | 0.42 | 0.36 | 0.30 | 0.74 | 0.65 | 0.85 | 0.85 | 0.31 | 0.40 | 0.26 | 0.35 | 0.67 | 0.68 |
|  | Ratio | 1.14 | | 1.19 | | 1.15 | | 1.00 | | 0.80 | | 0.72 | | 0.98 | |
| H & NHW | mean | 0.48 | 0.42 | 0.11 | 0.30 | 0.40 | 0.64 | 0.85 | 0.85 | 1.42 | 0.40 | 0.60 | 0.36 | 0.78 | 0.68 |
|  | Ratio | 1.16 | | 0.37 | | 0.62 | | 0.99 | | 3.57 | | 1.69 | | 1.15 | |
| **Predictor set identified from Lin et al.** | | | | | | | | | | | | | | | |
|  |  | PPV | | FPR | | TPR | | NPV | | te | | FNR | | ACC | |
|  |  | protected | privileged | protected | privileged | protected | privileged | protected | privileged | protected | privileged | protected | privileged | protected | privileged |
| NHB & NHW | mean | 0.48 | 0.43 | 0.31 | 0.29 | 0.67 | 0.65 | 0.82 | 0.86 | 0.47 | 0.40 | 0.33 | 0.35 | 0.68 | 0.69 |
|  | Ratio | 1.13 | | 1.07 | | 1.02 | | 0.96 | | 1.18 | | 0.96 | | 0.98 | |
| H & NHW | mean | 0.36 | 0.43 | 0.22 | 0.29 | 0.48 | 0.65 | 0.85 | 0.86 | 0.64 | 0.40 | 0.52 | 0.35 | 0.72 | 0.69 |
|  | Ratio | 0.85 | | 0.74 | | 0.73 | | 0.99 | | 1.60 | | 1.52 | | 1.04 | |

H = Hispanic; NHB = Non-Hispanic Black; NHW = Non-Hispanic White

**Table S8. Number of patients and their distribution by high versus low data-continuity in racial subgroups in REACHnet–LABlue**

| EHR data-continuity | NHW | NHB | Hispanic | Unknown | Others |
| --- | --- | --- | --- | --- | --- |
| High N = 7494 | 315 (4.2%) | 1317 (17.57%) | 5639 (75.25%) | 148 (1.97%) | 75 (1%) |
| Low N = 7073 | 176 (2.49%) | 939 (13.28%) | 5769 (81.56%) | 103 (1.46%) | 86 (1.22%) |

NHB: non-Hispanic Blacks, NHW; non-Hispanic Whites

**Table S9. Comparative performance of the models in REACHnet–LABlue.**

| Data | Model | AUROC | F1-Score | Specificity | Accuracy |
| --- | --- | --- | --- | --- | --- |
| Complete predictor set | XGBoost | 0.74 | 0.67 | 0.71 | 0.68 |
|  | LR | 0.76 | 0.72 | 0.41 | 0.65 |
| Predictor set (Huang et al.) | XGBoost | 0.65 | 0.54 | 0.76 | 0.60 |
|  | LR | 0.76 | 0.70 | 0.65 | 0.69 |
| Predictor set (Lin et al.) | XGBoost | 0.81 | 0.75 | 0.71 | 0.74 |
|  | LR | 0.78 | 0.73 | 0.38 | 0.65 |

*AUROC: area under the receiver operating characteristic

**Figure S1. Number of patients by HEPS score category in the (a) OneFlorida+ cohort, (b) REACHnet–LABlue cohort**

| (a)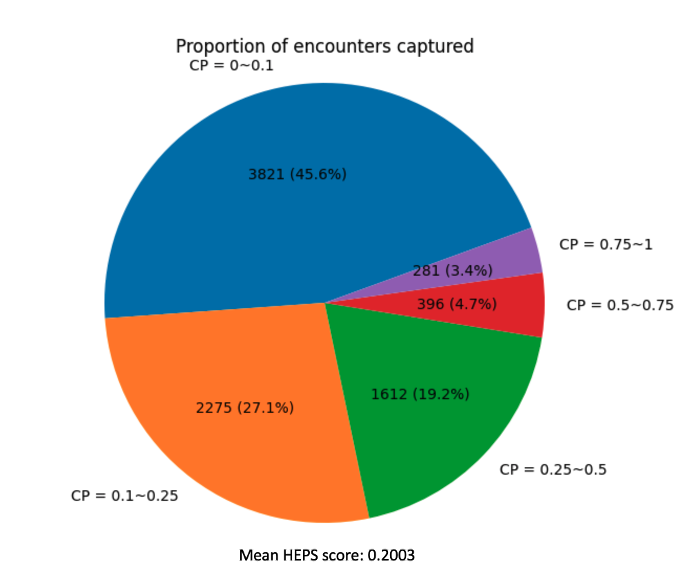 | (b)  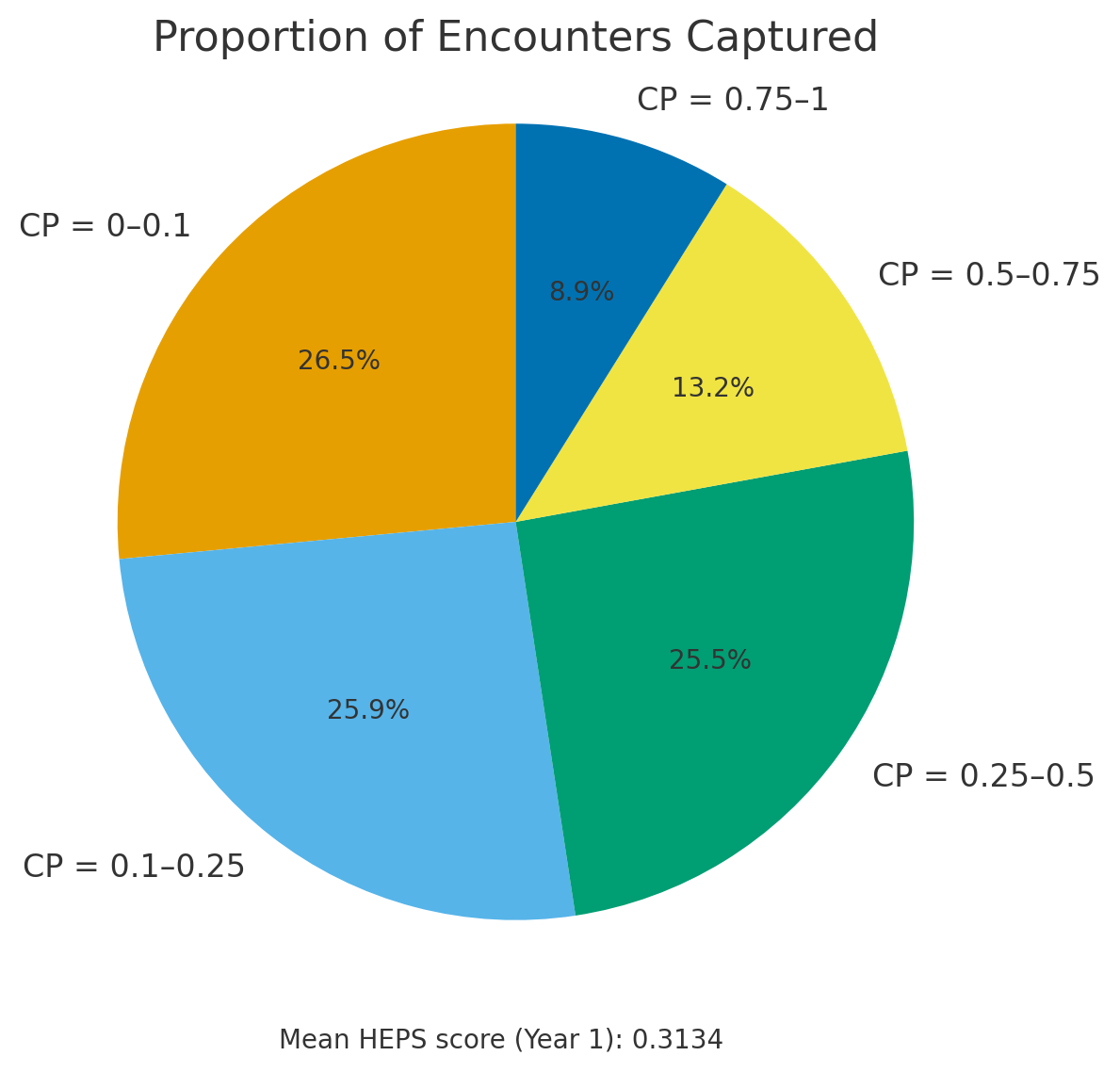 |
| --- | --- |

**Figure S2. Sensitivity of capturing clinical information in relation of EHR-continuity in OneFlorida+**

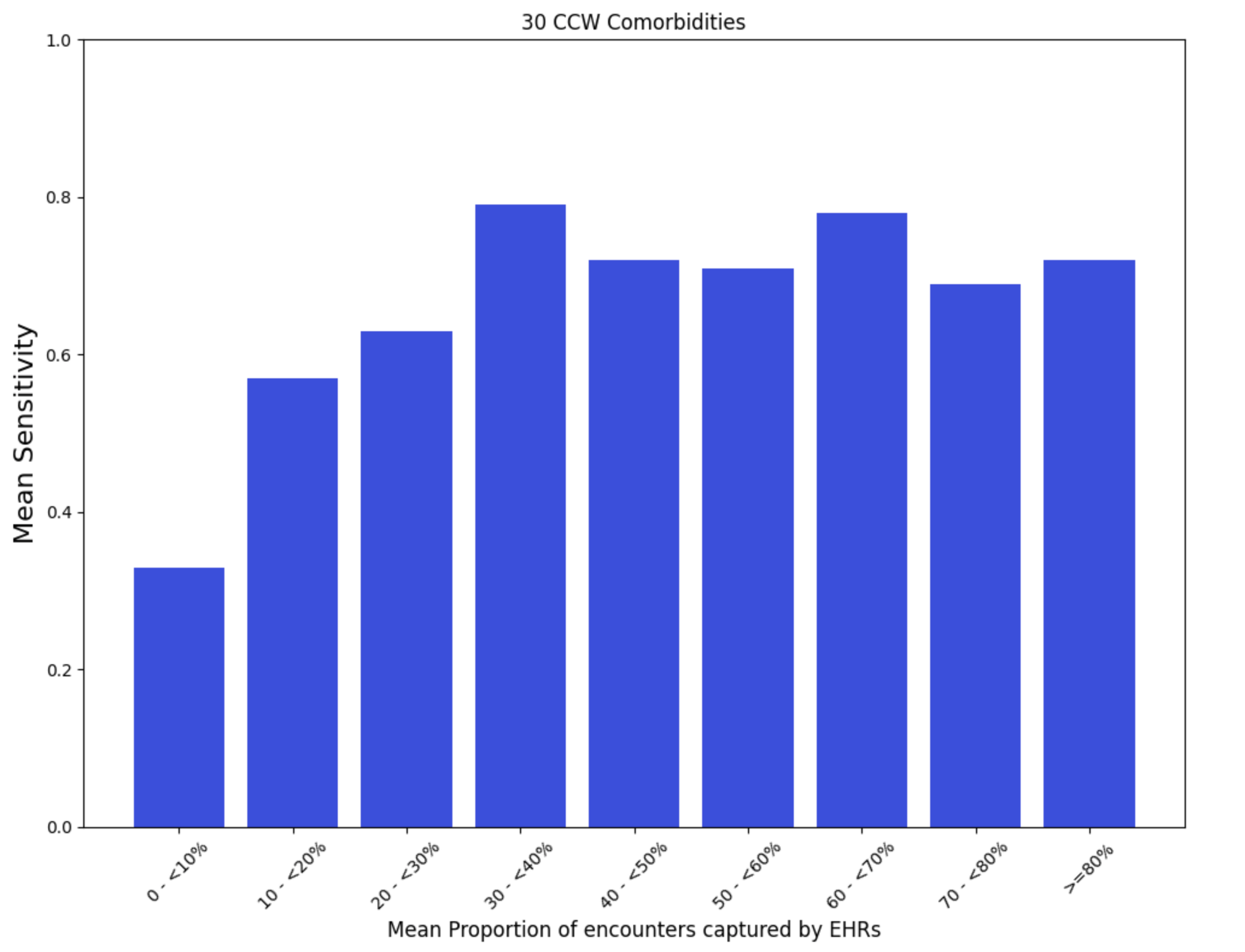

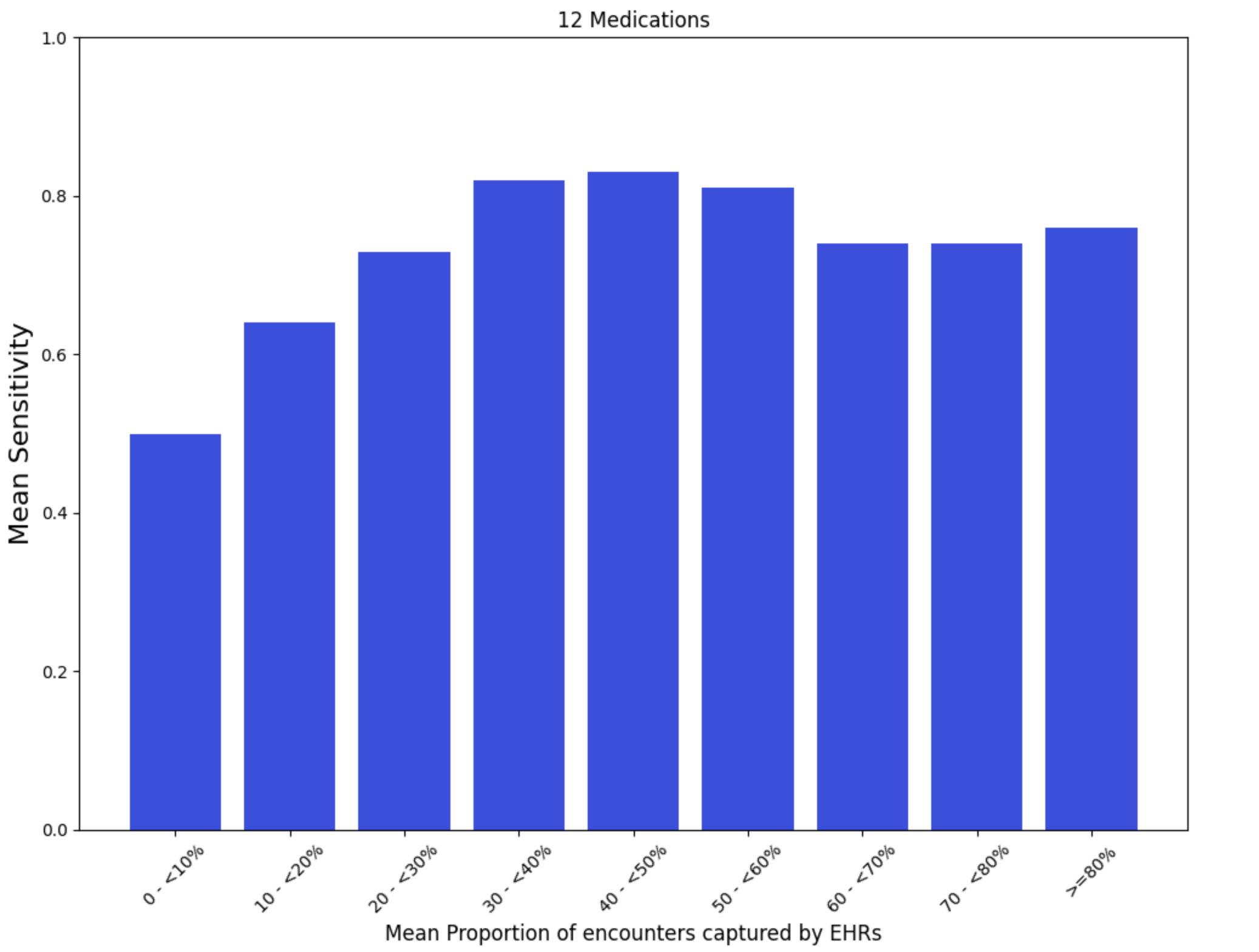

**Figure S3. SHAP values for XGBoost on the predictor set identified from Huang et al.**

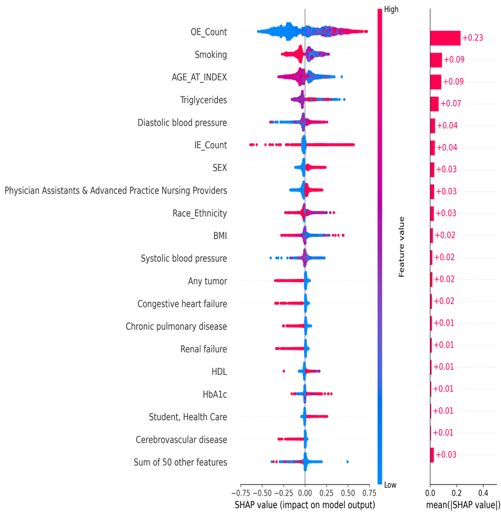

**Figure S4. SHAP values for XGBoost on the predictor set identified from Lin et al.**

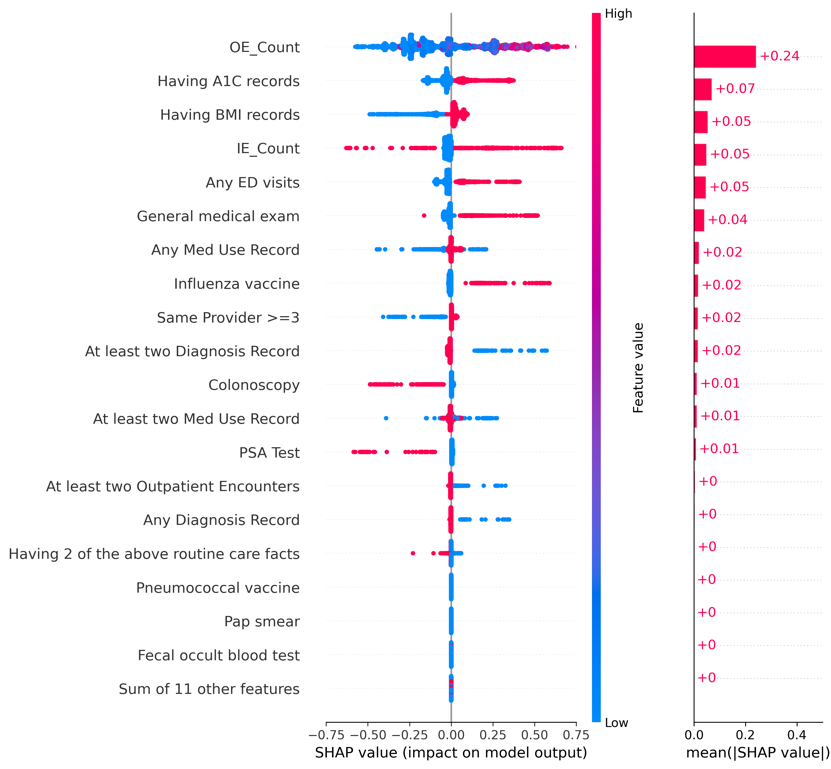

**Figure S5. SHAP values for Logistic Regression on the complete predictor set.**

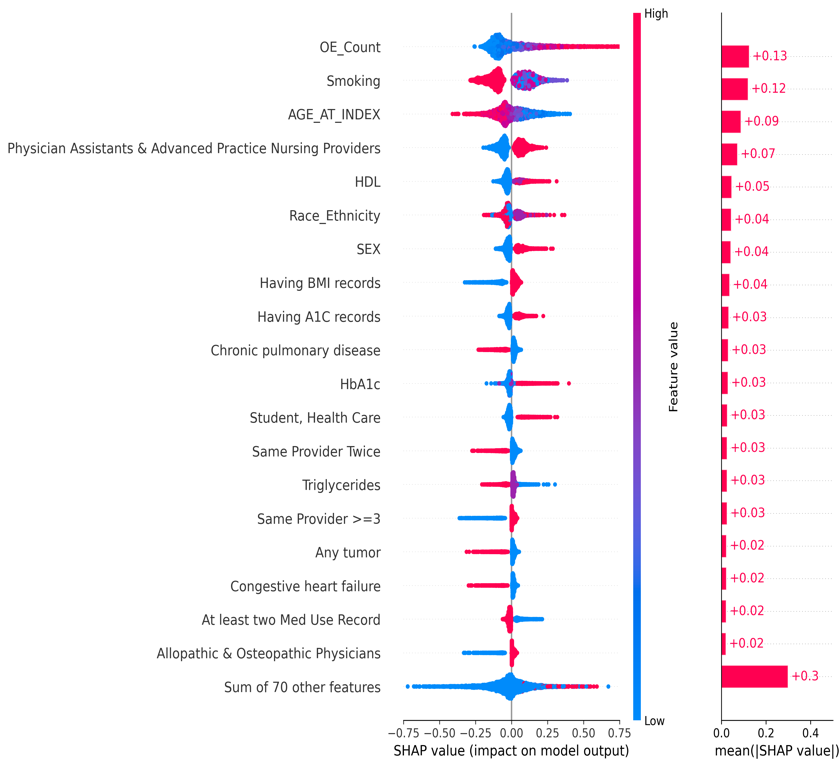

**Figure S6. SHAP values for Logistic Regression on the predictor set identified from Huang et al.**

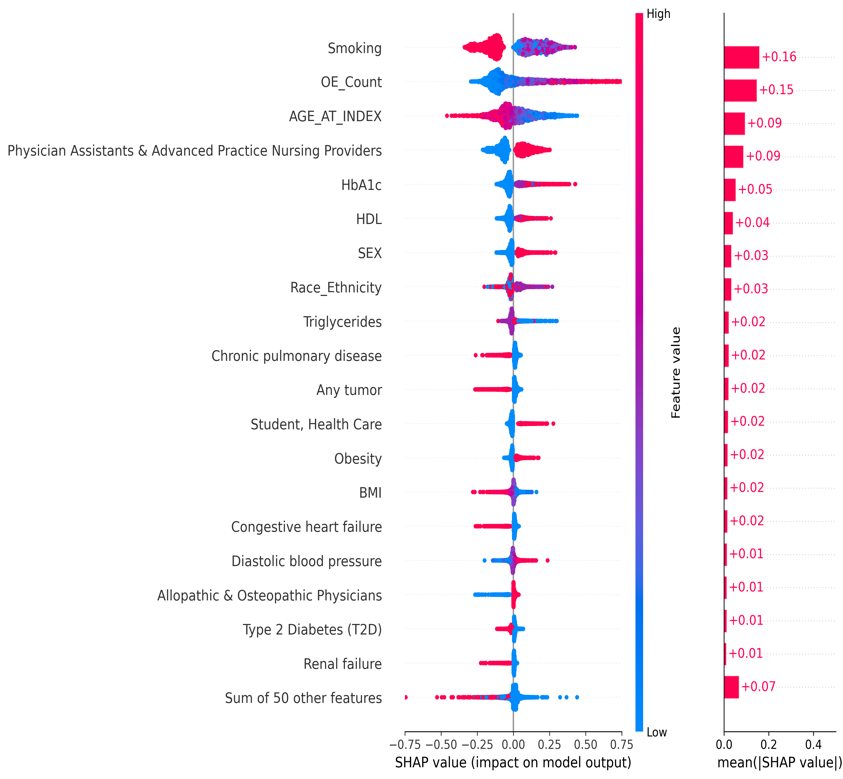

**Figure S7. SHAP values for Logistic Regression on the predictor set identified from Lin et al.**

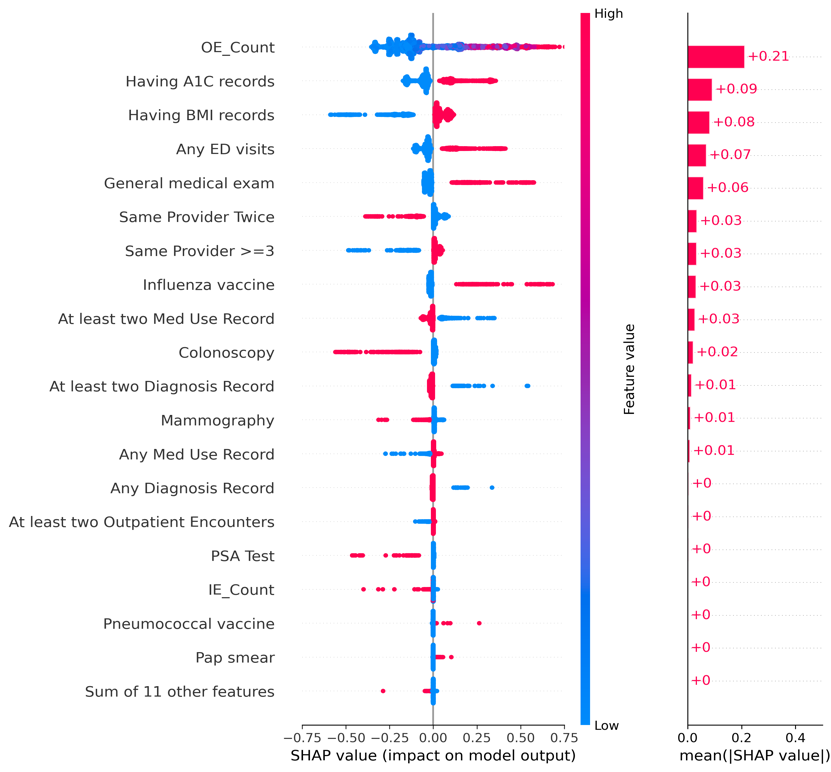

**Figure S8. Mean difference by HEPS score deciles in (a) CCI score, (b) ECI score, (c) Combined score in REACHnet–LABlue**

| (a) | (b) | (c) |
| --- | --- | --- |
| 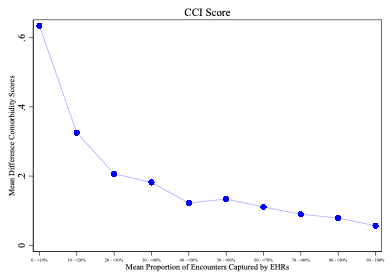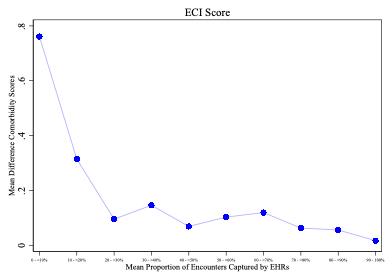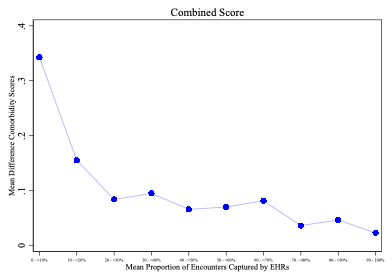 | | |

CCI = Charlson Comorbidity Index; ECI = Elixhauser Comorbidity Index

**Figure S9. Mean standardized difference between EHR-claims linked vs EHR-only data for (a) comorbidities and (b) medications in REACHnet–LABlue.**

| (a) | (b) |
| --- | --- |
| 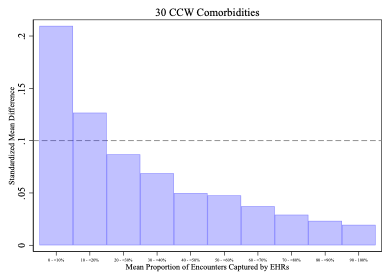 | 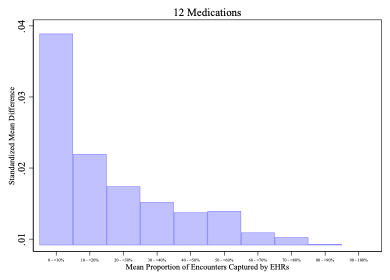 |
